# Associations of antibodies against several infections with Alzheimer disease neuropathology: a prospective cohort study analysis

**DOI:** 10.64898/2026.03.04.26347593

**Authors:** Caterina Felici, Rebecca E Green, Charlotte Warren-Gash, Julia Butt, Tim Waterboer, Alun D Hughes, Nishi Chaturvedi, Ashvini Keshavan, William Coath, Jonathan M Schott, Marcus Richards, Dylan M Williams

## Abstract

**Background and Objectives:** Associations of common infections with Alzheimer disease (AD) risk have been reported. A hypothesized mechanism to explain these is cerebral amyloid-beta (Aβ) aggregation as a defence in response to infection, with subsequent tau accumulation. However, few studies have assessed associations of infections with tau and Aβ pathology. We investigated associations of serological measures of several common infections with plasma p-tau217 and Aβ status measured by neuroimaging in the 1946 British birth cohort.

**Methods:** Circulating antibodies against 14 pathogens, measured at age 60-64 years, were modelled as pathogen serostatus (indicating lifetime exposure to an agent), pathogen burden indices (measuring cumulative exposure to 2+ pathogens), and seroreactivity tertiles (indicating recent immunological activity against pathogens). Associations of these were tested with plasma p-tau217 (primary outcome) and Aβ status measured by positron emission tomography imaging (Aβ-PET; secondary outcome), measured approximately 7 years after serology measurements. Modelling used multivariable quantile and logistic regression, respectively. Model 1 adjusted for sex and ages at serology and outcome assessment, models 2 and 3 additionally adjusted for *APOE* ε4 carriage and education, respectively. We also tested for interactions in associations with *APOE* ε4 carriage and education, and for interactions between herpes simplex virus 1 (HSV1) exposure with both cytomegalovirus (CMV) and varicella zoster virus (VZV) exposure.

**Results:** 1356 and 424 individuals had complete data for p-tau217 and Aβ-PET analyses, respectively. Mean age at p-tau217 was 69.9 years (SD 0.7) and 51.3% of participants were female. No notable associations were observed for either outcome in main models, with the exception being an unexpected relationship between seropositivity for herpes simplex virus 2 and lower p-tau217 at the 75th quantile. There was also some evidence for potential interactions in p-tau217 associations by *APOE* ε4 carriage (for *Helicobacter pylori* and CMV) and by educational attainment (for *Helicobacter pylori* serostatus).

**Discussion:** These findings are not supportive of associations between exposures to many common infections and aggregation of core AD neuropathology measures. The possibility that some pathogens might interact with *APOE* ε4 carriage and education in relation to AD neuropathology warrants further study.

## Introduction

There is increasing interest in potential roles of infections in dementia risk, particularly in relation to Alzheimer disease (AD)^1^. Suggested mechanisms underlying potential pathogenic effects include: i) the “antimicrobial protection hypothesis”, proposing that pathogens in the central nervous system trigger aggregation of Aβ peptides as a defensive mechanism; or ii) elicitation of a systemic inflammatory response to infections, which could facilitate the neuroinflammatory cascade observed in AD^2^. There might also be a greater role of overall infection burden rather than individual infectious agents in the development of dementia^3^. For instance, evidence suggests that varicella zoster virus (VZV) could be related to AD risk by enhancing reactivation of herpes simplex virus 1 (HSV1)^4^. An interaction has also been observed between HSV1 and cytomegalovirus (CMV) infection in relation to AD risk^5^. Since exposure to most herpesviruses and many other pathogens is common, yet only a subset of individuals develops clinical dementia, other factors might modify the relationship of pathogens with dementia. Some studies have reported that associations between common infections and dementia or cognition are more evident in individuals carrying *APOE* ε4 alleles ^6–8^― (the strongest genetic risk factor for late-onset AD) ―and in those with lower education^9,10^. However, there is scant evidence available on the association of common infections ― measured individually and in aggregate ― with core subclinical AD neuropathology (Aβ and phosphorylated tau deposition), nor for potential modifiers of any such associations^11–14^.

In this study, we examined prospective associations of evidence of exposure to 14 pathogens, individually and cumulatively, with measures of cerebral amyloidosis and tauopathy in a population-based birth cohort from the UK. We hypothesized that pathogen exposures would be associated with greater AD neuropathology, and that associations would be more prominent in individuals carrying one or two *APOE* ε4 alleles and in individuals with lower education.

## Methods

### Study population and design

The MRC National Survey of Health and Development (NSHD; also known as the 1946 British birth cohort) is a population-based birth cohort that enrolled 5,362 individuals born across mainland Britain during one week in March 1946, using stratified sampling on social class^15^. Participants have been followed continuously for 79 years to date. A subset of 502 NSHD participants was recruited into Insight 46, a neuroimaging sub-study with initial assessments at ages 69-71 years to collect neuropsychological, neuroimaging, and blood biomarkers for dementia^16^. This study principally uses data from: i) multiplex serology assays of plasma collected at ages 60-64 years in NSHD; ii) phosphorylated tau at threonine 217 (p-tau217) measures from plasma collected at home visits and clinical assessments at ages 69-71 in NSHD; iii) amyloid-β PET scans on individuals aged 69-71 years for the subset of participants in Insight 46 (Figure 1).

**Figure 1.**
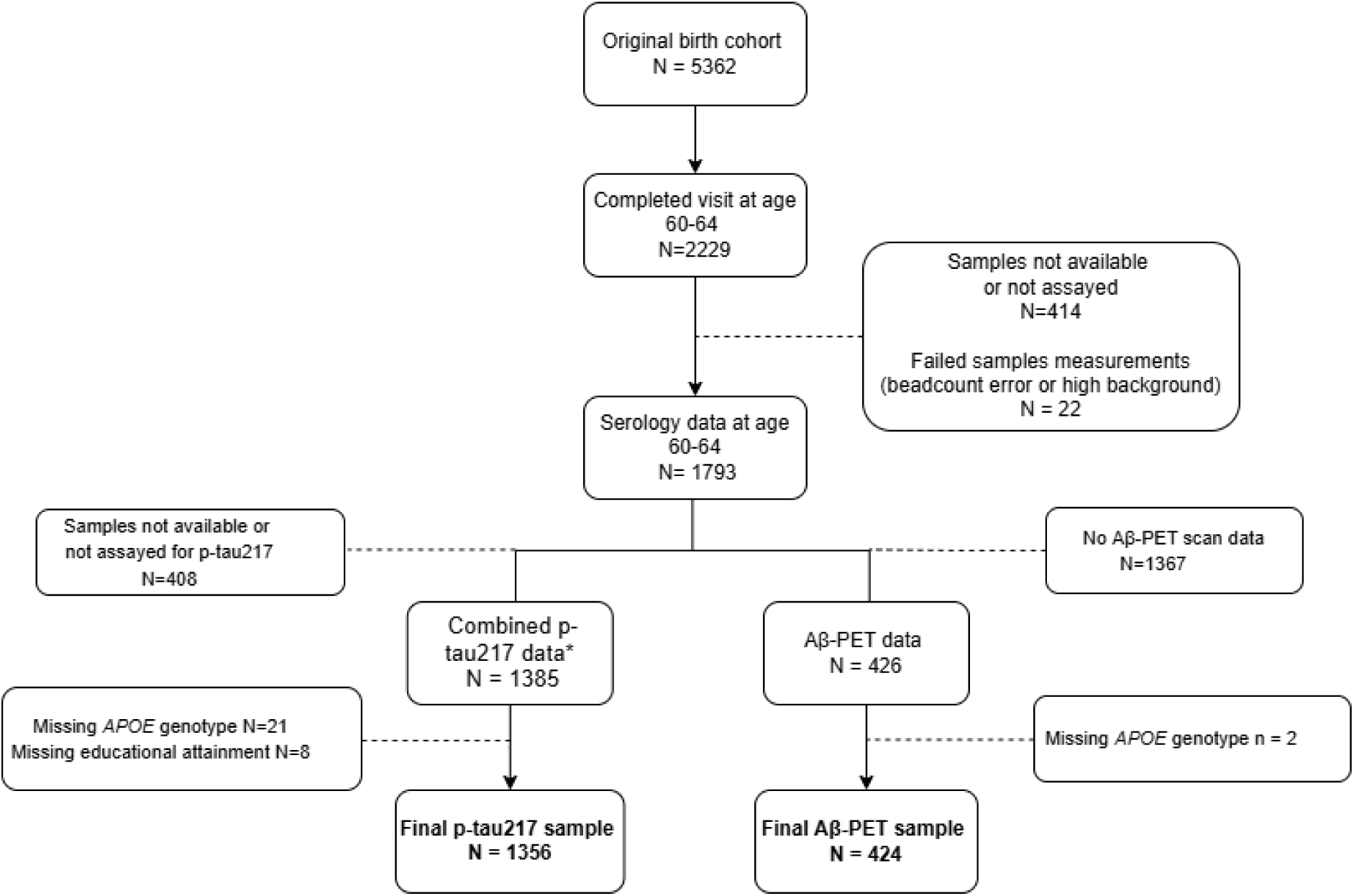
Study flowchart. Plasma p-tau217* from the NSHD home visits at age 69 (N = 952) were combined with p-tau217 data from Insight 46 participants (N = 484; ages 69-71) – see Supplemental Notes for more information.

### Assessment of pathogen exposures with multiplex serology

Exposure to pathogens was ascertained using a multiplex serology panel that quantifies circulating IgG antibody levels against a large number of antigens for different infectious agents, developed at the German Cancer Research Center (DKFZ) in Heidelberg^17,18^. We assayed 1813 plasma samples collected between ages 60 and 64 years (2006–2010)^19^. 1793 participants had valid multiplex serology data for 18 pathogens after quality control (Figure 1).

Serological values were used to define: 1) serostatus for individual pathogens, measured as a binary variable (seropositive/negative), indicating the presence or absence of detectable pathogen-specific antibodies and reflecting exposure at some point over the life course; 2) pathogen burden indices (PBIs) calculated as cumulative counts of serostatus variables; 3) seroreactivity tertiles derived from the continuum of antibody concentrations, where higher concentrations are thought to reflect more recent infection or re-exposure, or more frequent reactivation of latent pathogens. Serostatus for each pathogen was derived using thresholds for a specific antigen or a combination of different antigens (eTable 1). Two PBIs were derived: an index measuring the total number of infections and a neurotropic PBI restricted to a count of seropositive values for known neurotropic pathogens only (n=10, denoted with an asterisk in eTables 1 and 2), which might be more relevant to AD pathogenesis.

For seroreactivity analyses, we included seropositive participants and only addressed antigens with at least 30 observations in each tertile to avoid very small sample sizes. This resulted in the exclusion of all antigens for herpes simplex virus 2 (HSV2), *Toxoplasma gondii (*Tg), *Helicobacter pylori (*Hp), *Chlamydia trachomatis (*Ct) and some antigens for Human herpesvirus 6A (HHV-6A) and 6B (HHV-6B) in analyses using Aβ-PET as an outcome, while one antigen for HHV-6A was excluded in analyses of p-tau217. We excluded *Human papillomaviruses* and Kaposi’s sarcoma-associated herpesvirus (*KSHV)* from all analyses because of low seropositivity (< 5%). Risk of outcomes in higher vs. lowest tertiles of antibody levels were compared in statistical models.

### P-tau217 and Aβ-PET measurements

Plasma p-tau217 was the primary outcome, given the large sample size available with these data and its high accuracy for detecting AD neuropathology measured by PET neuroimaging^20^. This was assayed for home visit samples (N=952) and for samples collected at clinical visits by Insight 46 participants (N=484) using the ALZpath Simoa assay (Quanterix; absolute quantification yielding measurements in pg/mL)^20^. The assaying was undertaken at the UK DRI Biomarker Factory at UCL in separate batches for home visit and Insight 46 samples, with bridging between the batches. Further details are provided in the Supplemental Notes.

Cerebral Aβ deposition, the secondary outcome, was measured by PET (Aβ-PET) using the 18F-Florbetapir PET ligand and quantified via standardized uptake volume ratio (SUVR), with whole cerebellum used as a reference region following partial volume correction^16^. SUVR values were converted to the Centiloid scale^21^ [0 Centiloid = mean SUVR in controls; 100 Centiloid = mean SUVR in AD]. The value of 10 Centiloids was used as a threshold for amyloid positivity, considering a consensus view that cerebral amyloidosis can reliably be ruled out below 10 Centiloids^22^. Aβ-PET reconstruction was performed using the pseudo-CT method^16^. Pseudo-CT reconstructed values were imputed from the console-reconstructed values for 26 participants due to technical issues.

### Covariates

Covariates included age at serology, age at p-tau217 measurement or age at neuroimaging, sex, *APOE* genotype, and highest educational attainment. *APOE* and educational attainment are both associated with AD risk and possibly with different susceptibility to infections^23,24^. *APOE* genotypes were derived using variants rs7412 and rs429358 genotyped/imputed from genome-wide microarray data. A binary variable for *APOE* ε4 carriage was coded as: ε4 carriers (ε4/ε4, ε3/ε4, ε2/ε4) and ε4 non-carriers (ε2/ε3, ε3/ε3 or ε2/ε2). A binary variable categorized educational attainment as ordinary (O-level or equivalent or below) and advanced (A-Level or tertiary), based on self-report up to age 43 years.

## Statistical analysis

### Main analyses

Since this is an exploratory study, multiple testing correction was not performed. Complete-case analysis was performed since missing data (primarily for serology, p-tau and amyloid values) were unlikely to be amenable to multiple imputation (and the extent of these missing data was limited; Figure 1). All statistical tests were two-tailed at a significance level of 0.05. Analyses were performed in Stata 18 (StataCorp LLC, College Station, TX, USA). Figures were generated in R, version 4.3.2.

### Plasma p-tau217 modelling

Associations of serological measures with p-tau217 were examined using quantile regression at the median and the 75^th^ quantile. Quantile regression is robust to distributional assumptions and outliers. It also allows for testing of exposure-outcome associations at different parts of the outcome distribution, which may differ (for example, being more prominent at the tails) – especially for skewed outcomes, such as p-tau217 in this sample. Higher levels of p-tau217 are more likely to reflect higher levels of both amyloid and phosphorylated tau deposition.

We conducted four sets of quantile regression models. Model 1 adjusted for sex, age at serology, and age at p-tau217 measurement. Model 2 adjusted for model 1 covariates plus *APOE* ε4 carriage. Model 3 adjusted for model 1 covariates plus education. All statistical models included sampling weights to adjust for the initial sampling strategy in the NSHD (individuals from manual social class families were under-represented originally)^25^. For seroreactivity analyses, tertiles were modelled as an ordinal variable and orthogonal polynomial contrasts were applied to test for linear trends.

Two-way interaction terms were applied to model 1 to examine whether *APOE* ε4 carriage and education modify associations of serological measures with p-tau217. Additionally, separate regression models incorporating interaction terms were used to test whether infection by VZV and CMV modify the association of HSV1 serostatus with p-tau217, given previous suggestions of these interactions. Interactions were considered suggestive for *P* values less than 0.1. Interactions were tested only in median quantile regression. In analyses using seroreactivities tertiles as exposures, all interaction terms were tested simultaneously through a joint Wald test.

### Aβ-PET modelling

Secondary analyses followed the same methodology described above, with the following adaptations: a) Associations of serological measures with Aβ-PET positivity were examined via multivariable logistic regression; b) Model 1 was adjusted for sex, age at serology, and age at neuroimaging; c) interaction analyses were omitted because of reduced statistical power in the smaller sample. Linearity of continuous variable (ages at serology and neuroimaging) associations with log-odds of Aβ-PET positivity was assessed using fractional polynomials and quadratic terms, respectively, and indicated no requirements for non-linear modelling.

### Sensitivity analyses

In p-tau217 modelling, we conducted three sensitivity analyses. First, we repeated analyses without applying sampling weights. Second, we tested the effects of further adjusting analyses for BMI and estimated glomerular filtration rate (eGFR) to reduce noise in p-tau217 measurements, since body size and kidney function may influence variability in p-tau217 concentration^26,27^. Details of eGFR estimation are available in supplementary notes. Third, interaction terms were added to all models to assess whether relationships between serological measures and p-tau217 differed by source of p-tau measurements (home visits or attendance at Insight 46).

For Aβ-PET modelling, we repeated analyses: (1) without applying sampling weights; (2) excluding individuals with imputed data for Aβ-PET (N=22); (3) using a continuous measure of Centiloids as an outcome. Since Centiloids follow a gamma distribution, generalised linear models with a gamma log link were used. Centiloid values were adjusted to ensure positivity by adding the minimum value to all observations. Regression coefficients were multiplied by 100 to obtain relative percentage differences in the outcome per unit of exposure for ease of interpretation^28^.

To give context to association magnitudes from our modelling of the primary outcome, we also modelled associations of *APOE* ε4 carriage with p-tau217 with quantile regression at the median and 75^th^ quantile, adjusting for age at p-tau217 measurement and sex.

### Standard Protocol Approvals, Registrations, and Patient Consents

Approvals for the relevant NSHD follow-ups were granted from the Central Manchester Research Ethics Committee, the Scotland A Research Ethics Committee, and the Queen Square Research Ethics Committee in London, UK (references 07/H1008/245, 08/MRE00/12, 14/SS/1009, 14/LO/1073 and 14/LO/1173). Participants gave informed, written consent for all follow-ups.

## Results

### Sample characteristics

There were 1356 participants with complete data for p-tau217 analyses and 424 for Aβ-PET analyses (Figure 1). Table 1 shows characteristics for both analytical samples.

**Table 1.** Characteristics of each analytical sample.

|  | P-tau217<br>sample<br>(N=1,356) | Negative<br>(N=304, 71.7%) | Aβ-PET sample *<br>Positive<br>(N=120, 28.3%) | Total<br>(N=424) |
| --- | --- | --- | --- | --- |
| Age at serology, mean (SD) | 63.2 (1.1) | 63.4 (1.1) | 63.3 (1.1) | 63.4 (1.1) |
| Age at p-tau217, mean (SD) | 69.9 (0.7) |  |  |  |
| Age at neuroimaging, mean (SD) |  | 70.7 (0.7) | 70.6 (0.7) | 70.6 (0.7) |
| Sex, n (%) |  |  |  |  |
| Male | 661 (48.7%) | 155 (51.0%) | 64 (53.3%) | 219 (51.7%) |
| Female | 695 (51.3%) | 149 (49.0%) | 56 (46.7%) | 205 (48.3%) |
| APOE ε4 carriage, n (%) |  |  |  |  |
| non-carriers | 945 (69.7%) | 246 (80.9%) | 57 (47.5%) | 303 (71.5%) |
| carriers | 411 (30.3%) | 58 (19.1%) | 63 (52.5%) | 121 (28.5%) |
| Highest educational attainment (up to age 43 years), n (%) |  |  |  |  |
| Vocational or O-levels/equivalent or below | 698 (51.5%) | 126 (41.4%) | 43 (35.8%) | 169 (39.9%) |
| A-levels or higher | 658 (48.5%) | 178 (58.6%) | 77 (64.2%) | 255 (60.1%) |
| p-tau217, median [IQR] | 0.30 [0.21;0.44] |  |  |  |
| Centiloids, median [IQR] |  | -1.12 [-5.31; 1.98] | 33.64 [19.08; 66.47] | 1.00 [-3.50;14.76] |

Abbreviations: APOE = apolipoprotein E ; IQR = interquartile range; SD = standard deviation; NSHD = National Survey of Health and Development; Aβ = β-amyloid.
\* Aβ status defined by Centiloid 10 cut-off
Note: Age was measured in years and p-tau217 in pg/mL

Table 2 shows seroprevalences and PBIs for the analytical samples for p-tau217 and Aβ-PET analyses. Seroprevalences and PBIs for the larger sample of NSHD (and all of Insight 46) with serology data are available in eTable 2. There were no notable differences in serological variables in the analytical samples versus all with serology data available. Seroprevalences and sample characteristics are displayed separately for the two sub-samples comprising the p-tau217 analytical sample in eTables 3 and 4. Compared to the subset with p-tau data based on home visits at 69 years, the subset of participants with p-tau217 data based on the Insight 46 assessment were slightly older (as expected), had higher proportions of males and more educated participants, and had slightly lower p-tau217. Serological data were very similar between these subsets in all instances except MC virus serostatus (seropositivity was 8% higher in absolute terms in Insight 46).

**Table 2.**
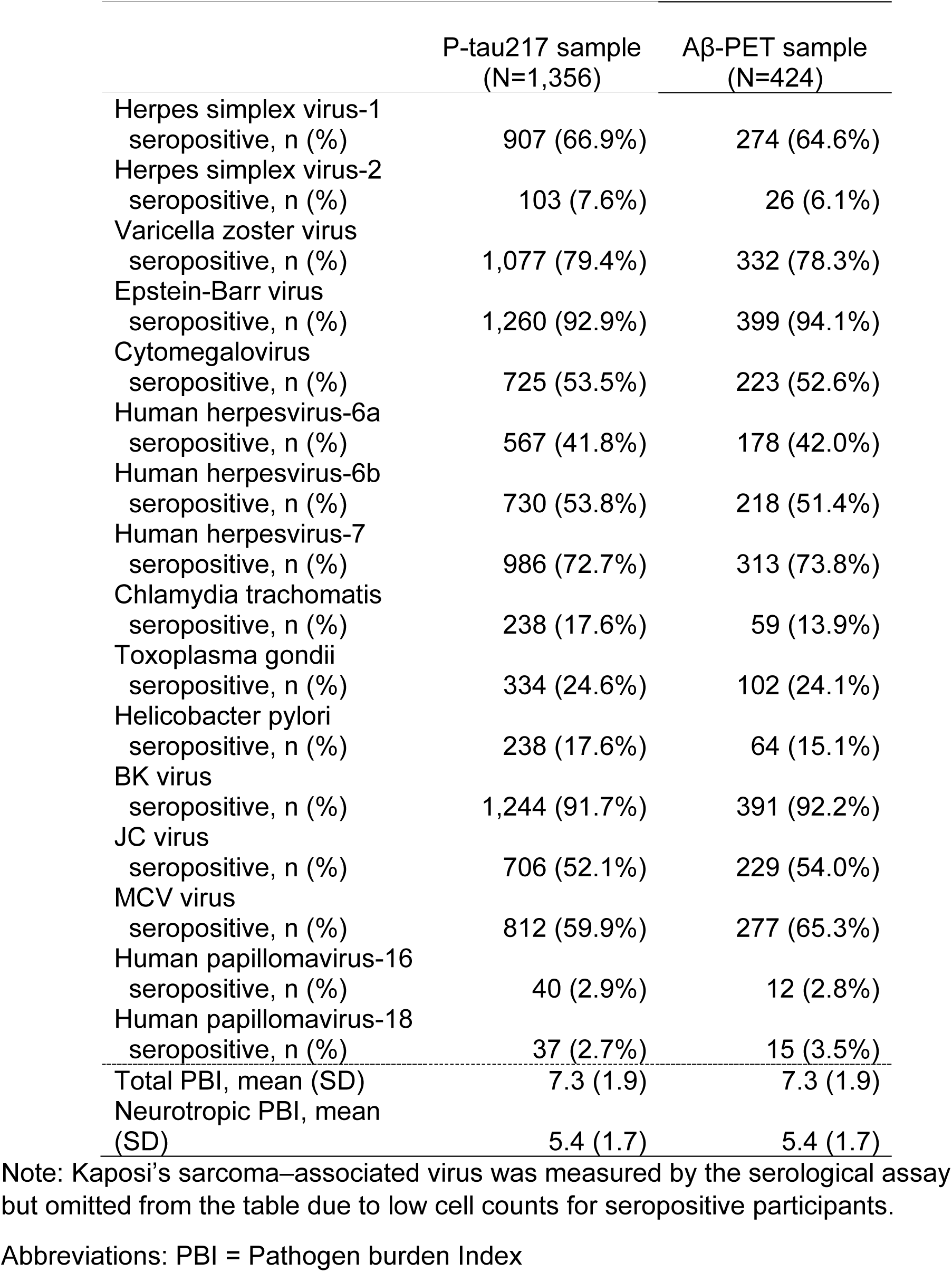
Seroprevalence in the analytical samples.

### Plasma p-tau217

Results for the associations of pathogen serostatus and pathogen burden with p-tau217 are plotted in Figure 2 and presented numerically in eTable 5.

**Figure 2.**
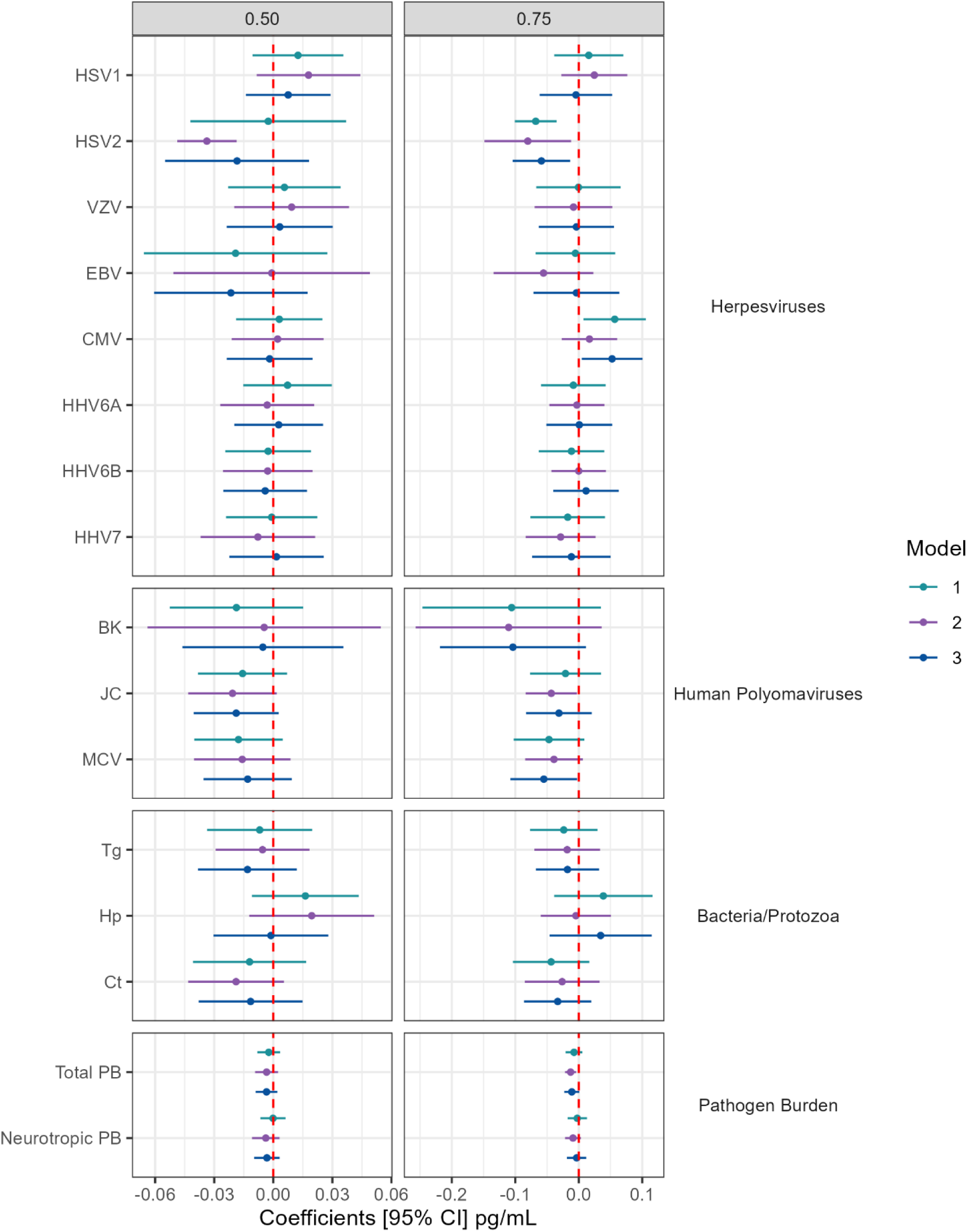
Associations of pathogen serostatus and pathogen burden with plasma p-tau217. Forest plots show weighted quantile regression results depicting differences (in mg/ml) in p-tau217 concentrations at the 50^th^ (i.e. median) and 75^th^ quantiles of the p-tau distribution per unit of exposure (left and right panels, respectively). Exposures were seropositive with reference to seronegative for individual pathogens, or per one additional pathogen in the pathogen burden indices. Model 1 adjusted for sex, age at serology, and age at p-tau217 measurement. Model 2 adjusted for model 1 covariates plus *APOE* ε4 carriage. Model 3 adjusted for model 1 covariates plus education. Abbreviations: HSV1 = Herpes simplex virus-1; HSV2 = Herpes simplex virus-2; VZV = Varicella zoster virus; EBV = Epstein-Barr virus; CMV = Cytomegalovirus; HHV6A = Human herpesvirus-6a; HHV6B = Human herpesvirus-6b; HHV7 = Human herpesvirus-7; BK = BK virus; JC = JC virus; MCV = Merkel cell polyomavirus; Tg = *Toxoplasma gondii*; Hp = *Helicobacter pylori*; Ct = *Chlamydia trachomatis*.

For most pathogens, serostatus was not notably or consistently associated with higher concentrations of p-tau217 at either the median or 75^th^ quantile of the p-tau distribution across model series. In most instances, p-tau differences by serostatus were modest, and confidence intervals included the null. For a comparison of magnitudes, in this sample, *APOE* ε4 carriage was associated with 0.09 pg/mL higher p-tau217 (95% CI: 0.05, 0.12) at the median, and 0.19 pg/mL higher p-tau217 (95% CI: 0.12, 0.26) at the 75^th^ quantile (eTable 6). The largest positive point estimate for any pathogen serostatus at the median of the p-tau distribution was 0.02 pg/mL for Hp (Model 1: 95% CI: -0.01, 0.04), and the highest at the 75^th^ quantile was 0.06 pg/mL for CMV (Model 1: 95% CI: 0.01, 0.11). As this indicates, there was evidence for p-tau217 concentration at the 75^th^ quantile being higher in individuals seropositive for CMV. However, this association magnitude was modest compared to *APOE* ε4’s effect on p-tau217 concentration at the 75^th^ quantile, and the CMV-p-tau217 association also attenuated when adjusted for *APOE* ε4 carriage. Seropositivity for HSV2 was associated with *lower* p-tau217 concentration at the 75^th^ quantile, while at the median only in Model 2 (adjusted for *APOE* ε4 carriage). Results were close to the null for both overall and neurotropic PBIs.

Associations of seroreactivities with p-tau217 are shown in eFigures 1-2 and eTable 7. Since there was no evidence for linear trends in seroreactivities tertiles for most antigens, these were modelled as ordinal variables. When examining associations with median p-tau217, lower p-tau217 concentrations were observed in those with the highest tertile for HHV6 IE1A (associations attenuated slightly after adjustment for ε4 carriage), for HP 887/2 (VacA-C) intermediate tertile when adjusted for ε4 carriage and education, while higher p-tau217 concentration were observed for the intermediate tertile for JC VP1. At the 75^th^ quantile, the EBV EAD intermediate tertile was associated with lower p-tau217 concentration, while the HHV6 IE1B highest tertile was associated with higher p-tau217 concentration but attenuated when adjusted for ε4 carriage. There were indications of association of the EBV EBNA peptide’s highest tertile with lower p-tau217 concentration at 75^th^ quantile, but these attenuated when adjusted for education.

### Interactions

We observed no interactions of HSV1 serostatus with either VZV or CMV serostatus in their associations with p-tau217 (eTable 8). There was suggestive evidence that associations of median p-tau217 with CMV and Hp serostatus, total pathogen burden, and CMV pp28 and VZV gE/gI antigens seroreactivities differed by *APOE* ε4 carriage. Suggestions for interactions by educational attainment was observed for Hp serostatus and some seroreactivity values for EBV, Hp and MC virus (eTable 9 and 10). Stratified results for *APOE* ε4 carriage are shown in eTable 11. Median p-tau217 concentration was higher for individuals seropositive for CMV (by 0.08 pg/mL, 95% CI: 0.01, 0.14) and Hp (by 0.08 pg/mL, 95% CI: 0.01, 0.16) among *APOE* ε4 carriers only. Total PBI was associated with nominally lower median p-tau217 in ε4 non-carriers only. Antigen CMV pp28 was associated with higher median p-tau217 concentration among *APOE* ε4 carriers only. On the other hand, VZV gE/gI was associated with lower concentration of median p-tau217 at both tertiles in ε4 carriers only. Stratified results for education are presented in eTable 12. Hp serostatus was associated with opposing differences in median p-tau217 in those with lower education (0.04 pg/mL higher p-tau217; 95% CI: 0.01, 0.07) and higher education (-0.05 pg/mL p-tau217; 95% CI: - 0.09, -0.01).

### Aβ-PET

Associations of pathogen serostatus and pathogen burden with odds of a positive Aβ-PET scan are presented in eFigure 3 and eTable 13. There was a suggestion of higher odds of Aβ-PET positivity for individuals seropositive for EBV (Model 1: OR: 2.84; 95% CI: 0.95, 8.52) and Tg (Model 1: OR:1.69; 95% CI: 0.94, 3.05), with a large point estimate, although considering the size of CIs, associations were estimated with low precision. To put association magnitudes in context, carriage of at least one ε4 allele associated with an OR of 5.16 for Aβ-PET positivity (95% CI 3.02, 8.81) in this cohort^29^. There was no evidence for associations of either PBIs nor seroreactivity tertiles with Aβ-PET positivity (eFigure 3-4 and eTables 13-14).

### Sensitivity analyses for plasma p-tau217

In unweighted analyses, results resembled the main findings, albeit with minor influence on evidence of interactions (primarily by education). Analyses further adjusted for BMI and eGFR also produced similar results (N = 1331).

When examining whether p-tau217 results differed by subsets of p-tau data (collected at home visits or Insight 46), there was evidence for interaction between source of p-tau data and HSV2 in relation to median p-tau217 in all three models and particularly when adjusted for *APOE* (*P* for interaction in model 2 <0.01). In stratified analyses, HSV2 showed evidence for association with lower median p-tau217 in home visit data only in model 2 (-0.06 pg/mL; 95% CI:-0.08, -0.04), with a point estimate in the opposite direction in Insight 46 data (0.03 pg/mL; 95% CI: -0.01, 0.08). There was a similar pattern of associations of *Tg* with p-tau217 by p-tau data collection source, albeit with less pronounced evidence for interaction (*P*=0.10 in model 1).

### Sensitivity analyses for Aβ-PET

In unweighted analyses, most results resembled the main analyses except for EBV and Tg serostatus findings, which were both less precise and smaller in magnitude. Results for pathogen serostatus, pathogen burden and seroreactivities did not change notably when excluding participants with imputed Aβ-PET. There was evidence for higher levels of Aβ deposition among EBV seropositives when repeating the analyses using a continuous variable for Centiloids (Model 1: 25% higher burden; 95% CI: 0, 51%). Results for the associations of PBIs and seroreactivities with Aβ deposition also resembled the main analysis.

Data from all sensitivity analyses are available on request.

## Discussion

In this population-based cohort from the UK, we observed no marked evidence for associations of exposure to many common pathogens, measured by antibody levels, with AD neuropathology at ages 69-71 years. There were some suggestive findings when interactions by *APOE* ε4 carriage and educational attainment were considered – though these may have arisen by chance.

HSV1 has been investigated extensively in relation to AD, including in terms of potential interactions with *APOE* ε4 carriage^6,30,31^. Our null findings are consistent with observations from some other epidemiological studies. One found associations of symptomatic herpesvirus infections with elevated plasma glial fibrillary acidic protein (GFAP), a marker of astrogliosis, but not with plasma Aβ42/40 or neurofilament light chain (NfL)^12^. Other previous studies reported similarly null results for serological measures of HSV and other common infections in relation to plasma Aβ^13,14^. We previously found null or little evidence for associations of most pathogens under investigation here (including HSV1) with MRI-derived measures of dementia risk, including hippocampal volume^19^. Another cohort study observed no associations of HSV, *Chlamydia pneumoniae*, Hp, and CMV serostatus with dementia incidence^32^.

There could be several reasons for null findings. First, the pathogens studied here may not be notable determinants of AD pathogenesis (at least as measured by the aggregation of AD’s cardinal proteinopathies by age ∼70). Alternatively, defining infection exposure from IgG antibodies does not provide a measure of infection severity: it is possible that severe infections might influence AD pathogenesis more than milder ones. Another caveat is that infections could be involved in later stages of disease development after AD pathophysiology is established, rather than in preclinical phases ^33,34^. With AD neuropathology assessment at ∼70 years in this study, some participants with developing AD may not have been far along the disease trajectory. Last, exposure to several pathogens and interactions among them might be necessary for the development of neuropathology. However, despite previously reported associations of PBIs with dementia risk^11^, we found no associations of PBIs or some key hypothesized pathogen interactions (HSV1 by VZV or CMV) with AD neuropathology measures here. We note that we would not have had sufficient statistical power to investigate the numerous possible two-way (or higher order) interactions between other combinations of individual pathogens.

Among *APOE* ε4 carriers only, we observed higher median p-tau217 concentration in individuals seropositive for CMV and with higher antibody levels against CMV pp28 antigen, and in individuals seropositive for Hp. A greater risk of dementia in ε4 carriers infected by HSV1 and other pathogens (HIV, SARS-CoV-2) has been observed elsewhere^23^. One study reported that ε4 carriers had higher levels of CMV IgG antibodies^35^. There is conflicting evidence from studies that have examined CMV and Hp in relation to outcomes other than those in the present analysis (dementia, cognition and neuroimaging), with evidence for and against interaction by ε4 carriage among these ^32,19,36^. Whether there might be a specific mechanism linking CMV or Hp to increased AD pathology in ε4 carriers alone, or whether these results might have arisen by chance, is unclear.

Some of the findings – including potential associations of higher pathogen burden with lower median p-tau217 among ε4 non-carriers and lower p-tau217 among ε4 carriers with high antibodies against VZV antigen gE/gl– were unexpected, and existing research in relation to these is scant and conflicting^37,38^. Our observation of lower p-tau217 at the 75^th^ quantile for individuals seropositive to HSV2 was also unanticipated and the scant evidence related to this association to date would imply no AD risk difference or increased risk according to HSV2 exposure^19,39^. Some antigen seroreactivities seemed to be variably and inconsistently associated with either higher or lower p-tau217 concentrations. Lastly, there was a suggestion of positive associations between Hp serostatus and median p-tau217 concentrations for lower levels of education, while negative associations for higher levels of education (individuals with lower education seem to be, on average, more exposed to or susceptible to infections overall, and to Hp infection specifically)^9,40,41,42^. Each of these suggestive findings could be due to chance – particularly given the degree of multiple testing performed here – or from biases such as residual confounding.

In Aβ-PET analyses, our findings suggested the possibility of associations of EBV and *Tg* serostatus with Aβ deposition. Contrasting findings between serological measures of infections with measures of amyloidosis and tauopathy have been observed elsewhere, where higher antibody levels for EBV EBNA, CMV and HSV1 associated with lower Aβ_42/40_ (reflecting higher cortical amyloid) but to lower pTau-181 (reflecting lower amyloid-induced tauopathy)^43^. However, given the smaller sample size for Aβ-PET analysis and the lack of these associations with p-tau217 (which is a strong predictor of Aβ status), these findings may not be robust.

### Strengths and limitations

This study has several strengths. We measured exposure to many common infections via serology (i.e. not via self-reporting of clinically-manifest infections) alongside large-scale data on accurate and specific biomarkers of AD pathology^20^. Measurement of AD pathology with these biomarkers allows the study of AD neuropathology in a preclinical phase. The use of subclinical biomarkers as outcomes rather than a clinical endpoint for AD may reduce the influence of some biases, such as reverse causation and survival bias.

There are also several limitations. As an observational study, residual confounding can affect association estimates, though for this to explain our mostly null findings, we would need to have unaccounted factors strong enough to mask more prominent associations from being observed. NSHD participants with poorer health, worse socioeconomic circumstances and lower childhood cognitive ability were under-represented in the 68–69 year and Insight 46 assessments, producing the possibility of selection bias^44,45^. If infections had increased study attrition due to heighted morbidity or mortality between exposure and outcome measurement, these exposures could also have impacted selection. However, most of the infections being studied do not have common clinical manifestations among individuals aged 60-70 years and we are unaware of data showing major increases in mortality risk for seropositive individuals in this age range^46^. The lack of longitudinal data for serological markers and knowledge of exact timing of infection did not allow temporal characterisation of exposures and increases the risk of exposure misclassification. Although we studied numerous infections – many of which have been implicated in AD aetiology previously – some with existing evidence of association with AD could not be addressed by measurement with the current multiplex serology platform, e.g. *Chlamydia pneumoniae* and periodontal pathogens. Finally, since the NSHD is comprised of white British participants, generalizability to other populations may be limited.

### Future directions

Though these findings cast doubt on the roles of many common infections in the development of AD neuropathology, future research in this area could help to establish whether more fine-grained characterization of infections (including timing, dose and severity) or other infections not studied here are relevant to AD pathogenesis. Studying infections in relation to other biomarkers of neurodegenerative and neurovascular diseases including inflammatory responses could help to clarify potential effects of pathogens in other causes of dementia besides AD. Finally, the generation of similar exposure data in large cohorts involving older participants with appreciable dementia incidence would help to test associations of pathogens individually and in combination with dementia risk directly. Other associations could be pertinent, considering strong evidence that suggests shingles vaccination (and potentially other vaccines) reduces dementia incidence^47^, though not necessarily via an effect on AD pathogenesis specifically.

## Acknowledgments

We are grateful to study participants of the NSHD (and Insight 46). We are also grateful to the radiographers and nuclear medicine physicians at the UCL Institute of Nuclear Medicine, to the staff at the Leonard Wolfson Experimental Neurology Centre at UCL and to Dr Carole Sudre (Unit for Lifelong Health and Ageing, UCL) and Dr Michalis Katsoulis (Unit for Lifelong Health and Ageing, UCL) for providing guidance for analyses.

## Data availability

NSHD data are available to researchers subject to approval by the cohort’s Data Sharing Committee. For details, visit: https://skylark.ucl.ac.uk/NSHD/

