## Supplemental material for "Associations of antibodies against several infections with Alzheimer disease neuropathology: a prospective cohort study analysis"

**Supplemental Notes**

*Plasma p-tau217 assaying*

Initial plasma p-tau217 assaying was undertaken using blood collected from Insight 46 participants at age ~70. Following this, a subset of samples available from the NSHD's 69-year home visits were selected for further assaying to prioritize: i) those without p-tau217 measures from Insight46; ii) individuals who had died following the 69-year visits and had no prospect of joining later Insight 46 follow-ups (where further p-tau217 assaying is planned). The selection of the remainder of the available 69-year home visit samples chosen for assaying was at random. Approximately half of all available 69-year samples were assayed in total (insufficient funds were available for all samples).

Levels of p-tau217 were determined in both sets of plasma samples in singlicate using the ALZpath Simoa V2 assay (Quanterix),[1] and values were adjusted for cross-run variation within each sample set using pooled plasma control samples assayed on each run. As part of the assaying of p-tau217 on samples from age 69, 18 plasma samples previously assayed in singlicate in Insight 46 and 2 pooled plasma samples (also used in the previous Insight 46 assaying) were re-assayed in duplicate to allow for bridging of the NSHD data to the Insight 46 results.

*Deriving estimated glomerular filtration rate (eGFR)*

We estimated eGFR at 69 years using the 2021 Chronic Kidney Disease Epidemiology Collaboration (CKD-EPI) Creatinine Equation[2] :

$$\mathbf{eGFR = 142 \times \min(SCr/\kappa,1)^{\alpha} \times \max(SCr/\kappa,1)^{-1.200} \times 0.9938^{Age} \times 1.012\ [if\ female]}$$

Abbreviations/units

- eGFR = estimated GFR in mL/min/1.73 $m^2$
- SCr = standardized serum creatinine in mg/dL
- κ = 0.7 (females) or 0.9 (males)
- α = -0.241 (females) or -0.302 (males)
- min = indicates the minimum of SCr/κ or 1
- max = indicates the maximum of SCr/κ or 1
- age = years

Evidence suggests using both creatinine and cystatin to estimate eGFR[3], however, since cystatin at 69 years was not available, we opted for the equation including only creatinine.

We pooled creatinine values from NSHD and Insight 46. Measures were in different units (mmol/L vs umol/L) and we converted them all to mmol/L[(umol/L)/1000]. We then converted creatinine values from mmol/L to mg/dl to be used in the equation described above using the following conversion:

Creatinine (mg/dL)=Creatinine (mmol/L)×11.3122

*References*

1. Ashton NJ, Brum WS, Di Molfetta G, et al. Diagnostic accuracy of a plasma phosphorylated tau 217 immunoassay for Alzheimer disease pathology. JAMA Neurol. 2024;81(3):255–263.
2. eGFR Equations for Adults - NIDDK. National Institute of Diabetes and Digestive and Kidney Diseases. Accessed September 9, 2025. https://www.niddk.nih.gov/research-funding/research-programs/kidney-clinical-research-epidemiology/laboratory/glomerular-filtration-rate-equations/adults
3. Inker LA, Eneanya ND, Coresh J, et al. New Creatinine- and Cystatin C–Based Equations to Estimate GFR without Race. New England Journal of Medicine. 2021;385(19):1737-1749. doi:10.1056/NEJMoa2102953

**eTable 1. Pathogens addressed in this study, with respective antigens, serostatus definition details and thresholds.**

| **Pathogens** | **Antigens and use for defining serostatus** | **Thresholds for mean fluorescence intensity values used to indicate seropositivity **** |
|---|---|---|
| Herpes simplex virus-1 (HSV1) * | 1gG | 170 |
| Herpes simplex virus-2 (HSV2) * | 2mgG unique | 180 |
| Varicella zoster virus (VZV) * | gE_gI | 100 |
| Epstein Barr virus (EBV) * | ≥2 positive out of:<br>EBNA<br>EA-D<br>VCAp18<br>Zebra | <br>411<br>110<br>2526<br>74 |
| Cytomegalovirus (CMV) * | ≥2 positive out of:<br>pp150NTerm<br>pp52<br>pp28 | <br>100<br>854<br>100 |
| Human herpesvirus-6A (HHV6A) * | IE1A and/or<br>p100 | 100<br>75 |
| Human herpesvirus-6B (HHV6B) * | IE1B and/or<br>p101K | 100<br>100 |
| Human betaherpesvirus 7 (HHV7) * | U14 | 225 |
| Kaposi's sarcoma-associated virus (KSHV) | LANA 3 and<br>K8.1 | 100<br>100 |
| BK virus (BKV) | VP1 | 250 |
| JC virus (JCV) * | VP1 | 100 |
| Merkel Cell virus (MCV) | VP1 | 250 |
| Human papillomavirus-16 (HPV16) | L1 | 100 |
| Human papillomavirus-18 (HPV18) | L1 | 100 |
| *Toxoplasma gondii* * | p22 and/or<br>sag1 | 150<br>150 |

| | | |
|---|---|---|
| *H.pylori* | ≥3 positive out of 8:<br>HP 10 (GroEL)<br>HP1098 (Hcp C)<br>HP 1564 (OMP)<br>HP 887/2 (VacA-C)<br>HP547/1 (CagA-N)<br>HP305<br>HP73 (UreA)<br>HP875 (Catalase) | <br>100<br>200<br>400<br>250<br>400<br>150<br>250<br>400 |
| *Chlamydia trachomatis* | pGP3 | 95 |

* *Neurotropic pathogens*

** *Thresholds were supplied by the developers of the multiplex platform.*

**eTable 2: Seroprevalences among all individuals with serology data in the Insight-46 subsample and the whole NSHD sample**

| | Insight-46 (N=468) | | NSHD (N=1793) | |
|---|---|---|---|---|
| | **N** | **(%)** | **N** | **(%)** |
| Herpes simplex virus-1* serostatus | | | | |
| seropositive | 304 | 65.0 | 1206 | 67.3 |
| Herpes simplex virus-2* serostatus | | | | |
| seropositive | 28 | 6.0 | 131 | 7.3 |
| Varicella zoster virus* serostatus | | | | |
| seropositive | 366 | 78.2 | 1424 | 79.4 |
| Epstein-Barr virus* serostatus | | | | |
| seropositive | 441 | 94.2 | 1666 | 92.9 |
| Cytomegalovirus* serostatus | | | | |
| seropositive | 238 | 50.9 | 972 | 54.2 |
| Human herpesvirus-6a* serostatus | | | | |
| seropositive | 196 | 41.9 | 764 | 42.6 |
| Human herpesvirus-6b* serostatus | | | | |
| seropositive | 242 | 51.7 | 967 | 53.9 |
| Human herpesvirus-7* serostatus | | | | |
| seropositive | 345 | 73.7 | 1304 | 72.7 |
| BK virus serostatus | | | | |
| seropositive | 429 | 91.7 | 1639 | 91.4 |
| JC virus serostatus | | | | |
| seropositive | 250 | 53.4 | 926 | 51.6 |
| MCV virus serostatus | | | | |
| seropositive | 305 | 65.2 | 1070 | 59.7 |
| Human papillomavirus-16 serostatus | | | | |
| seropositive | 13 | 2.8 | 49 | 2.7 |
| Human papillomavirus-18 serostatus | | | | |
| seropositive | 16 | 3.4 | 43 | 2.4 |
| T.gondii* serostatus | | | | |
| seropositive | 113 | 24.1 | 438 | 24.4 |
| H.pylori serostatus | | | | |
| seropositive | 68 | 14.5 | 315 | 17.6 |
| C.trachomatis serostatus | | | | |
| seropositive | 63 | 13.5 | 321 | 17.9 |
| | **Mean** | **SD** | **Mean** | **SD** |
| Total PBI, mean (SD) | 7.2 | (1.9) | 7.3 | (1.9) |
| Neurotropic PBI, mean (SD) | 5.4 | (1.6) | 5.5 | (1.6) |

Abbreviations: PBI = Pathogen burden Index; SD = standard deviation; NSHD = National Survey of Health and Development

* *Neurotropic pathogens*

**eTable 3. Sample characteristics for the NSHD home visit and Insight 46 subsamples used for p-tau217 analyses**

| | NSHD (N=902) | Insight46 (N=454) | Total (N=1,356) | Test[a] |
|---|---|---|---|---|
| Age at serology, mean (SD) | 63.1 (1.1) | 63.3 (1.0) | 63.2 (1.1) | 0.001 |
| Age at p-tau217, mean (SD) | 69.5 (0.2) | 70.6 (0.7) | 69.9 (0.7) | <0.001 |
| Sex, n (%) | | | | 0.055 |
| Male | 423 (46.9%) | 238 (52.4%) | 661 (48.7%) | |
| Female | 479 (53.1%) | 216 (47.6%) | 695 (51.3%) | |
| APOE e4 carriage, n (%) | | | | 0.564 |
| e4 non-carriers | 624 (69.2%) | 321 (70.7%) | 945 (69.7%) | |
| e4 carriers | 278 (30.8%) | 133 (29.3%) | 411 (30.3%) | |
| Highest educational attainment (up to age 43 years), n (%) | | | | <0.001 |
| Vocational or O-levels/equivalent or below | 516 (57.2%) | 182 (40.1%) | 698 (51.5%) | |
| A-levels or higher | 386 (42.8%) | 272 (59.9%) | 658 (48.5%) | |
| p-tau 217, median [IQR] | 0.32 [0.23-0.46] | 0.25 [0.18-0.36] | 0.30 [0.21-0.44] | <0.001 |

Abbreviations: APOE = apolipoprotein E ; IQR = interquartile range; SD = standard deviation; NSHD = National Survey of Health and Development; Aβ = β-amyloid.

[a] Pearson's chi-squared test for categorical variables and pooled t-test for continuous variables, except for p-tau217, for which Kruskal–Wallis rank test was used, comparing the NSHD and Insight46 groups

**eTable 4. Seroprevalences and PBIs for the NSHD home visit and Insight 46 subsamples used for p-tau217 analyses**

| | NSHD (N=902) | Insight46 (N=454) | Total (N=1,356) | p-value[a] |
|---|---|---|---|---|
| Herpes simplex virus-1 | | | | |
| seropositive | 612 (67.8%) | 295 (65.0%) | 907 (66.9%) | 0.289 |
| Herpes simplex virus-2 | | | | |
| seropositive | 75 (8.3%) | 28 (6.2%) | 103 (7.6%) | 0.159 |
| Varicella zoster virus | | | | |
| seropositive | 723 (80.2%) | 354 (78.0%) | 1,077 (79.4%) | 0.348 |
| Epstein-Barr virus | | | | |
| seropositive | 832 (92.2%) | 428 (94.3%) | 1,260 (92.9%) | 0.168 |
| Cytomegalovirus | | | | |
| seropositive | 492 (54.5%) | 233 (51.3%) | 725 (53.5%) | 0.261 |
| Human herpesvirus-6a | | | | |
| seropositive | 380 (42.1%) | 187 (41.2%) | 567 (41.8%) | 0.741 |
| Human herpesvirus-6b | | | | |
| seropositive | 492 (54.5%) | 238 (52.4%) | 730 (53.8%) | 0.459 |
| Human herpesvirus-6 | | | | |
| seropositive | 594 (65.9%) | 284 (62.6%) | 878 (64.7%) | 0.230 |
| Human herpesvirus-7 | | | | |
| seropositive | 650 (72.1%) | 336 (74.0%) | 986 (72.7%) | 0.448 |
| C.trachomatis | | | | |
| seropositive | 175 (19.4%) | 63 (13.9%) | 238 (17.6%) | 0.012 |
| T.gondii | | | | |
| seropositive | 224 (24.8%) | 110 (24.2%) | 334 (24.6%) | 0.807 |
| H.pylori | | | | |
| seropositive | 171 (19.0%) | 67 (14.8%) | 238 (17.6%) | 0.055 |
| BK virus | | | | |
| seropositive | 826 (91.6%) | 418 (92.1%) | 1,244 (91.7%) | 0.754 |
| JC virus | | | | |
| seropositive | 460 (51.0%) | 246 (54.2%) | 706 (52.1%) | 0.268 |
| MCV virus | | | | |
| seropositive | 516 (57.2%) | 296 (65.2%) | 812 (59.9%) | 0.005 |
| Human papillomavirus-16 | | | | |
| seropositive | 27 (3.0%) | 13 (2.9%) | 40 (2.9%) | 0.894 |
| Human papillomavirus-18 | | | | |
| seropositive | 22 (2.4%) | 15 (3.3%) | 37 (2.7%) | 0.356 |
| Total PBI (Mean, sd) | 7.3 (1.9) | 7.3 (1.9) | 7.3 (1.9) | 0.463 |
| Neurotropic PBI (Mean, sd) | 5.5 (1.7) | 5.4 (1.6) | 5.4 (1.7) | 0.468 |

Note: Kaposi's sarcoma–associated virus is omitted because of low cell counts for seropositive participants.

Abbreviations: PBI = Pathogen burden Index

[a] Pearson's chi-squared test

**eTable 5. Associations of pathogen serostatus and pathogen burden with differences in plasma p-tau217 (at the median and 75th quantile of the p-tau distribution).**

| | | Model 1 | | | Model 2 | | | Model 3 | | |
|---|---|---|---|---|---|---|---|---|---|---|
| **Exposure** | **Quantile** | **Coef** | **95% CI** | **p-value** | **Coef** | **95% CI** | **p-value** | **Coef** | **95% CI** | **p-value** |
| HSV1 | 0.50 | 0.01 | (-0.01, 0.04) | 0.29 | 0.02 | (-0.01, 0.04) | 0.18 | 0.01 | (-0.01, 0.03) | 0.49 |
| HSV1 | 0.75 | 0.02 | (-0.04, 0.07) | 0.57 | 0.02 | (-0.03, 0.08) | 0.36 | 0.00 | (-0.06, 0.05) | 0.88 |
| HSV2 | 0.50 | 0.00 | (-0.04, 0.04) | 0.90 | -0.03 | (-0.05, -0.02) | <0.001 | -0.02 | (-0.05, 0.02) | 0.32 |
| HSV2 | 0.75 | -0.07 | (-0.10, -0.03) | <0.001 | -0.08 | (-0.15, -0.01) | 0.02 | -0.06 | (-0.10, -0.01) | 0.01 |
| VZV | 0.50 | 0.01 | (-0.02, 0.03) | 0.70 | 0.01 | (-0.02, 0.04) | 0.53 | 0.00 | (-0.02, 0.03) | 0.81 |
| VZV | 0.75 | 0.00 | (-0.07, 0.07) | 0.99 | -0.01 | (-0.07, 0.05) | 0.79 | 0.00 | (-0.06, 0.06) | 0.90 |
| EBV | 0.50 | -0.02 | (-0.07, 0.03) | 0.42 | 0.00 | (-0.05, 0.05) | 0.97 | -0.02 | (-0.06, 0.02) | 0.28 |
| EBV | 0.75 | -0.01 | (-0.07, 0.06) | 0.87 | -0.06 | (-0.13, 0.02) | 0.16 | 0.00 | (-0.07, 0.06) | 0.91 |
| CMV | 0.50 | 0.00 | (-0.02, 0.02) | 0.79 | 0.00 | (-0.02, 0.03) | 0.85 | 0.00 | (-0.02, 0.02) | 0.87 |
| CMV | 0.75 | 0.06 | (0.01, 0.11) | 0.02 | 0.02 | (-0.03, 0.06) | 0.45 | 0.05 | (0.00, 0.10) | 0.03 |
| HHV6A | 0.50 | 0.01 | (-0.02, 0.03) | 0.53 | 0.00 | (-0.03, 0.02) | 0.80 | 0.00 | (-0.02, 0.03) | 0.81 |
| HHV6A | 0.75 | -0.01 | (-0.06, 0.04) | 0.74 | 0.00 | (-0.05, 0.04) | 0.89 | 0.00 | (-0.05, 0.05) | 0.98 |
| HHV6B | 0.50 | 0.00 | (-0.02, 0.02) | 0.81 | 0.00 | (-0.03, 0.02) | 0.81 | 0.00 | (-0.03, 0.02) | 0.70 |
| HHV6B | 0.75 | -0.01 | (-0.06, 0.04) | 0.66 | 0.00 | (-0.04, 0.04) | 0.99 | 0.01 | (-0.04, 0.06) | 0.66 |
| HHV7 | 0.50 | 0.00 | (-0.02, 0.02) | 0.94 | -0.01 | (-0.04, 0.02) | 0.60 | 0.00 | (-0.02, 0.03) | 0.89 |
| HHV7 | 0.75 | -0.02 | (-0.08, 0.04) | 0.56 | -0.03 | (-0.08, 0.03) | 0.31 | -0.01 | (-0.07, 0.05) | 0.71 |
| BK | 0.50 | -0.02 | (-0.05, 0.02) | 0.28 | 0.00 | (-0.06, 0.05) | 0.88 | -0.01 | (-0.05, 0.04) | 0.80 |
| BK | 0.75 | -0.11 | (-0.25, 0.03) | 0.14 | -0.11 | (-0.26, 0.04) | 0.14 | -0.10 | (-0.22, 0.01) | 0.08 |
| JC | 0.50 | -0.02 | (-0.04, 0.01) | 0.18 | -0.02 | (-0.04, 0.00) | 0.07 | -0.02 | (-0.04, 0.00) | 0.09 |
| JC | 0.75 | -0.02 | (-0.08, 0.03) | 0.46 | -0.04 | (-0.08, 0.00) | 0.04 | -0.03 | (-0.08, 0.02) | 0.24 |
| MCV | 0.50 | -0.02 | (-0.04, 0.00) | 0.12 | -0.02 | (-0.04, 0.01) | 0.21 | -0.01 | (-0.04, 0.01) | 0.26 |
| MCV | 0.75 | -0.05 | (-0.10, 0.01) | 0.10 | -0.04 | (-0.08, 0.01) | 0.09 | -0.06 | (-0.11, 0.00) | 0.04 |
| Tg | 0.50 | -0.01 | (-0.03, 0.02) | 0.61 | -0.01 | (-0.03, 0.02) | 0.65 | -0.01 | (-0.04, 0.01) | 0.31 |
| Tg | 0.75 | -0.02 | (-0.08, 0.03) | 0.38 | -0.02 | (-0.07, 0.03) | 0.49 | -0.02 | (-0.07, 0.03) | 0.48 |
| Hp | 0.50 | 0.02 | (-0.01, 0.04) | 0.24 | 0.02 | (-0.01, 0.05) | 0.23 | 0.00 | (-0.03, 0.03) | 0.94 |
| Hp | 0.75 | 0.04 | (-0.04, 0.12) | 0.33 | 0.00 | (-0.06, 0.05) | 0.87 | 0.03 | (-0.05, 0.12) | 0.40 |
| Ct | 0.50 | -0.01 | (-0.04, 0.02) | 0.41 | -0.02 | (-0.04, 0.01) | 0.13 | -0.01 | (-0.04, 0.01) | 0.39 |
| Ct | 0.75 | -0.04 | (-0.10, 0.02) | 0.16 | -0.03 | (-0.08, 0.03) | 0.38 | -0.03 | (-0.09, 0.02) | 0.22 |

#### eTable 5. Continued

| | | Model 1 | | | Model 2 | | | Model 3 | | |
|---|---|---|---|---|---|---|---|---|---|---|
| **Exposure** | **Quantile** | **Coef** | **95% CI** | **p-value** | **Coef** | **95% CI** | **p-value** | **Coef** | **95% CI** | **p-value** |
| Total PBI | 0.50 | 0.00 | (-0.01, 0.00) | 0.44 | 0.00 | (-0.01, 0.00) | 0.26 | 0.00 | (-0.01, 0.00) | 0.22 |
| Total PBI | 0.75 | -0.01 | (-0.02, 0.01) | 0.27 | -0.01 | (-0.02, 0.00) | 0.00 | -0.01 | (-0.02, 0.00) | 0.07 |
| Neurotropic PB | 0.50 | 0.00 | (-0.01, 0.01) | 0.96 | 0.00 | (-0.01, 0.00) | 0.28 | 0.00 | (-0.01, 0.00) | 0.33 |
| Neurotropic PB | 0.75 | 0.00 | (-0.02, 0.01) | 0.76 | -0.01 | (-0.02, 0.00) | 0.15 | 0.00 | (-0.02, 0.01) | 0.66 |

Note: quantile regression results depicting differences in p-tau217 concentrations at the $50^{th}$ and $75^{th}$ quantiles of the p-tau distribution per unit of exposure; seropositive with reference to seronegative for individual pathogens, or per 1 additional pathogen in the pathogen burden indices. Model 1 adjusted for sex, age at serology, and age at p-tau217 measurement. Model 2 adjusts for model 1 covariates plus *APOE* ε4 carriage. Model 3 adjusts for model 1 covariates plus education.

Abbreviations: HSV1 = Herpes simplex virus-1; HSV2 = Herpes simplex virus-2; VZV = Varicella zoster virus; EBV = Epstein-Barr virus; CMV = Cytomegalovirus; HHV6A = Human herpesvirus-6a; HHV6B = Human herpesvirus-6b; HHV7 = Human herpesvirus-7; BK = BK virus; JC = JC virus; MCV = Merkel cell polyomavirus; Tg = *Toxoplasma gondii*; Hp = *Helicobacter pylori*; Ct = *Chlamydia trachomatis;* PBI = Pathogen burden Index.

**eTable 6. Associations of APOE ε4 carriage with plasma p-tau217 (at the median and 75th quantile of the p-tau distribution)**

| | | Model 1 | | | Model 2 | | |
|---|---|---|---|---|---|---|---|
| **APOE ε4 carriage** | **Quantile** | **Coef** | **95% CI** | **p-value** | **Coef** | **95% CI** | **p-value** |
| Carriers | 0.50 | 0.08 | (0.04, 0.12) | <0.001 | 0.09 | (0.05, 0.12) | <0.001 |
| Carriers | 0.75 | 0.18 | (0.11, 0.24) | <0.001 | 0.19 | (0.12, 0.26) | <0.001 |

Note: Model 1: unadjusted; Model 2: adjusted for age at p-tau217 measurement and sex.

**eTable 7. Assocs of pathogen antigen seroreactivities with plasma p-tau217 (at the median and 75th quantile of the p-tau distribution).**

| | | | Model 1 | | | Model 2 | | | Model 3 | | | |
|---|---|---|---|---|---|---|---|---|---|---|---|---|
| **Pathogen** | **Tertiles** | **Quantile** | **Coef** | **95% CI** | **p-value** | **Coef** | **95% CI** | **p-value** | **Coef** | **95% CI** | **p-value** | **N** |
| HSV1_1gG | 2nd | 0.50 | -0.01 | (-0.05, 0.02) | 0.41 | -0.02 | (-0.05, 0.02) | 0.34 | -0.04 | (-0.07, -0.01) | 0.01 | 907 |
| | 3rd | 0.50 | 0.00 | (-0.03, 0.04) | 0.76 | 0.00 | (-0.03, 0.03) | 0.94 | -0.02 | (-0.05, 0.01) | 0.12 | 907 |
| HSV1_1gG | 2nd | 0.75 | 0.01 | (-0.08, 0.09) | 0.84 | -0.02 | (-0.09, 0.05) | 0.58 | 0.03 | (-0.05, 0.10) | 0.48 | 907 |
| | 3rd | 0.75 | -0.04 | (-0.11, 0.03) | 0.29 | -0.02 | (-0.09, 0.04) | 0.47 | -0.02 | (-0.08, 0.04) | 0.53 | 907 |
| HSV2_2mgGunique | 2nd | 0.50 | 0.01 | (-0.07, 0.08) | 0.90 | 0.01 | (-0.05, 0.08) | 0.68 | 0.01 | (-0.09, 0.10) | 0.90 | 103 |
| | 3rd | 0.50 | -0.01 | (-0.07, 0.04) | 0.66 | 0.00 | (-0.06, 0.05) | 0.86 | -0.01 | (-0.09, 0.06) | 0.70 | 103 |
| HSV2_2mgGunique | 2nd | 0.75 | 0.03 | (-0.13, 0.20) | 0.68 | 0.11 | (-0.07, 0.29) | 0.22 | 0.09 | (-0.09, 0.26) | 0.33 | 103 |
| | 3rd | 0.75 | -0.02 | (-0.13, 0.09) | 0.71 | 0.03 | (-0.09, 0.15) | 0.60 | -0.02 | (-0.16, 0.12) | 0.82 | 103 |
| VZV_gE_gI | 2nd | 0.50 | 0.00 | (-0.03, 0.03) | 0.97 | -0.01 | (-0.04, 0.02) | 0.50 | 0.01 | (-0.02, 0.04) | 0.43 | 1077 |
| | 3rd | 0.50 | 0.00 | (-0.03, 0.02) | 0.73 | -0.01 | (-0.04, 0.02) | 0.42 | 0.00 | (-0.03, 0.03) | 0.88 | 1077 |
| VZV_gE_gI | 2nd | 0.75 | 0.00 | (-0.08, 0.09) | 0.91 | 0.05 | (-0.01, 0.11) | 0.13 | 0.03 | (-0.06, 0.11) | 0.55 | 1077 |
| | 3rd | 0.75 | -0.05 | (-0.12, 0.03) | 0.21 | -0.03 | (-0.08, 0.02) | 0.24 | -0.02 | (-0.10, 0.05) | 0.59 | 1077 |
| EBV_VCAp18 | 2nd | 0.50 | 0.02 | (-0.01, 0.04) | 0.19 | 0.02 | (-0.01, 0.05) | 0.20 | 0.02 | (-0.01, 0.05) | 0.19 | 1222 |
| | 3rd | 0.50 | 0.01 | (-0.02, 0.03) | 0.56 | 0.02 | (-0.01, 0.05) | 0.29 | 0.01 | (-0.02, 0.03) | 0.47 | 1222 |
| EBV_VCAp18 | 2nd | 0.75 | 0.02 | (-0.06, 0.11) | 0.55 | 0.02 | (-0.04, 0.08) | 0.49 | 0.03 | (-0.04, 0.11) | 0.35 | 1222 |
| | 3rd | 0.75 | -0.01 | (-0.08, 0.05) | 0.69 | 0.00 | (-0.05, 0.05) | 0.93 | 0.01 | (-0.06, 0.07) | 0.88 | 1222 |
| EBV_EBNA_peptide | 2nd | 0.50 | -0.02 | (-0.05, 0.00) | 0.07 | -0.02 | (-0.05, 0.01) | 0.28 | -0.03 | (-0.06, 0.00) | 0.07 | 1170 |
| | 3rd | 0.50 | -0.03 | (-0.06, 0.00) | 0.10 | -0.04 | (-0.07, 0.00) | 0.03 | -0.02 | (-0.05, 0.01) | 0.17 | 1170 |
| EBV_EBNA_peptide | 2nd | 0.75 | -0.03 | (-0.12, 0.05) | 0.40 | -0.06 | (-0.13, 0.00) | 0.07 | -0.05 | (-0.13, 0.04) | 0.28 | 1170 |
| | 3rd | 0.75 | -0.08 | (-0.15, 0.00) | 0.04 | -0.06 | (-0.11, -0.01) | 0.03 | -0.06 | (-0.15, 0.02) | 0.12 | 1170 |
| EBV_Zebra | 2nd | 0.50 | 0.01 | (-0.02, 0.04) | 0.61 | 0.01 | (-0.02, 0.04) | 0.66 | 0.01 | (-0.02, 0.04) | 0.40 | 1201 |
| | 3rd | 0.50 | 0.00 | (-0.03, 0.03) | 0.86 | 0.01 | (-0.02, 0.04) | 0.61 | 0.00 | (-0.03, 0.03) | 0.97 | 1201 |
| EBV_Zebra | 2nd | 0.75 | -0.02 | (-0.11, 0.06) | 0.57 | 0.00 | (-0.05, 0.05) | 0.99 | 0.05 | (-0.03, 0.12) | 0.23 | 1201 |
| | 3rd | 0.75 | -0.02 | (-0.09, 0.05) | 0.54 | 0.00 | (-0.05, 0.05) | 0.90 | 0.00 | (-0.06, 0.07) | 0.95 | 1201 |
| EBV_EAD | 2nd | 0.50 | 0.01 | (-0.02, 0.04) | 0.58 | -0.01 | (-0.04, 0.02) | 0.57 | 0.01 | (-0.02, 0.04) | 0.48 | 1170 |
| | 3rd | 0.50 | 0.00 | (-0.04, 0.03) | 0.88 | -0.02 | (-0.05, 0.01) | 0.21 | 0.01 | (-0.03, 0.04) | 0.68 | 1170 |
| EBV_EAD | 2nd | 0.75 | -0.07 | (-0.13, -0.01) | 0.02 | -0.07 | (-0.11, -0.02) | <0.001 | -0.08 | (-0.14, -0.01) | 0.03 | 1170 |
| | 3rd | 0.75 | -0.05 | (-0.12, 0.02) | 0.16 | -0.04 | (-0.10, 0.01) | 0.11 | -0.06 | (-0.13, 0.02) | 0.12 | 1170 |
| CMV_pp150NTerm | 2nd | 0.50 | 0.02 | (-0.02, 0.07) | 0.33 | 0.02 | (-0.02, 0.06) | 0.32 | 0.03 | (-0.01, 0.06) | 0.19 | 750 |
| | 3rd | 0.50 | 0.00 | (-0.04, 0.05) | 0.93 | -0.01 | (-0.05, 0.03) | 0.72 | 0.00 | (-0.03, 0.04) | 0.92 | 750 |
| CMV_pp150NTerm | 2nd | 0.75 | -0.01 | (-0.12, 0.10) | 0.84 | 0.00 | (-0.08, 0.08) | 0.96 | -0.02 | (-0.12, 0.07) | 0.60 | 750 |
| | 3rd | 0.75 | -0.05 | (-0.16, 0.05) | 0.32 | -0.05 | (-0.14, 0.04) | 0.27 | -0.06 | (-0.15, 0.03) | 0.16 | 750 |

### eTable 7. Continued.

| | | | Model 1 | | | Model 2 | | | Model 3 | | | |
|---|---|---|---|---|---|---|---|---|---|---|---|---|
| **Pathogen** | **Tertiles** | **Quantile** | **Coef** | **95% CI** | **p-value** | **Coef** | **95% CI** | **p-value** | **Coef** | **95% CI** | **p-value** | **N** |
| CMV_pp52 | 2nd | 0.50 | -0.01 | (-0.05, 0.03) | 0.76 | -0.01 | (-0.05, 0.04) | 0.80 | -0.01 | (-0.05, 0.03) | 0.50 | 725 |
| | 3rd | 0.50 | 0.01 | (-0.03, 0.05) | 0.76 | 0.03 | (-0.01, 0.08) | 0.15 | 0.00 | (-0.04, 0.04) | 0.97 | 725 |
| CMV_pp52 | 2nd | 0.75 | 0.01 | (-0.11, 0.13) | 0.91 | 0.03 | (-0.05, 0.11) | 0.49 | 0.04 | (-0.06, 0.14) | 0.47 | 725 |
| | 3rd | 0.75 | 0.01 | (-0.09, 0.12) | 0.83 | 0.02 | (-0.05, 0.09) | 0.56 | 0.00 | (-0.10, 0.11) | 0.96 | 725 |
| CMV_pp28 | 2nd | 0.50 | 0.01 | (-0.02, 0.04) | 0.59 | 0.01 | (-0.02, 0.05) | 0.46 | 0.01 | (-0.03, 0.04) | 0.76 | 766 |
| | 3rd | 0.50 | 0.02 | (-0.02, 0.06) | 0.25 | 0.03 | (-0.01, 0.06) | 0.12 | 0.01 | (-0.03, 0.05) | 0.61 | 766 |
| CMV_pp28 | 2nd | 0.75 | -0.01 | (-0.13, 0.11) | 0.86 | -0.02 | (-0.10, 0.06) | 0.60 | -0.01 | (-0.13, 0.11) | 0.89 | 766 |
| | 3rd | 0.75 | -0.01 | (-0.10, 0.08) | 0.88 | 0.03 | (-0.06, 0.12) | 0.51 | -0.01 | (-0.10, 0.08) | 0.86 | 766 |
| HHV6_IE1A_truncated | 2nd | 0.50 | -0.02 | (-0.07, 0.03) | 0.53 | -0.04 | (-0.10, 0.01) | 0.11 | -0.01 | (-0.07, 0.04) | 0.59 | 521 |
| | 3rd | 0.50 | -0.05 | (-0.09, -0.01) | 0.01 | -0.05 | (-0.10, 0.00) | 0.05 | -0.05 | (-0.09, -0.01) | 0.01 | 521 |
| HHV6_IE1A_truncated | 2nd | 0.75 | -0.03 | (-0.16, 0.09) | 0.62 | -0.07 | (-0.17, 0.02) | 0.14 | -0.04 | (-0.17, 0.09) | 0.57 | 521 |
| | 3rd | 0.75 | -0.10 | (-0.23, 0.02) | 0.11 | -0.09 | (-0.17, -0.01) | 0.03 | -0.10 | (-0.23, 0.03) | 0.12 | 521 |
| HHV6_p100truncated | 2nd | 0.50 | 0.00 | (-0.11, 0.10) | 0.98 | 0.02 | (-0.09, 0.12) | 0.75 | 0.02 | (-0.08, 0.12) | 0.75 | 93 |
| | 3rd | 0.50 | -0.01 | (-0.07, 0.05) | 0.73 | 0.00 | (-0.06, 0.06) | 0.98 | 0.01 | (-0.05, 0.06) | 0.80 | 93 |
| HHV6_p100truncated | 2nd | 0.75 | 0.00 | (-0.26, 0.25) | 0.97 | 0.03 | (-0.11, 0.17) | 0.66 | 0.01 | (-0.26, 0.27) | 0.96 | 93 |
| | 3rd | 0.75 | -0.03 | (-0.24, 0.19) | 0.80 | -0.01 | (-0.13, 0.10) | 0.80 | -0.04 | (-0.26, 0.19) | 0.73 | 93 |
| HHV6_IE1B_truncated | 2nd | 0.50 | 0.03 | (-0.01, 0.07) | 0.16 | 0.00 | (-0.05, 0.04) | 0.87 | 0.02 | (-0.01, 0.06) | 0.21 | 663 |
| | 3rd | 0.50 | 0.02 | (-0.02, 0.06) | 0.44 | 0.02 | (-0.02, 0.06) | 0.34 | 0.03 | (-0.02, 0.07) | 0.22 | 663 |
| HHV6_IE1B_truncated | 2nd | 0.75 | 0.08 | (0.00, 0.17) | 0.06 | 0.04 | (-0.02, 0.10) | 0.21 | 0.08 | (0.01, 0.16) | 0.04 | 663 |
| | 3rd | 0.75 | 0.12 | (0.04, 0.20) | <0.001 | 0.05 | (-0.03, 0.14) | 0.21 | 0.12 | (0.04, 0.19) | <0.001 | 663 |
| HHV6_p101Ktruncated | 2nd | 0.50 | -0.03 | (-0.10, 0.04) | 0.39 | -0.01 | (-0.08, 0.05) | 0.72 | 0.00 | (-0.08, 0.07) | 0.92 | 167 |
| | 3rd | 0.50 | 0.02 | (-0.06, 0.10) | 0.63 | 0.02 | (-0.08, 0.11) | 0.76 | 0.02 | (-0.05, 0.09) | 0.50 | 167 |
| HHV6_p101Ktruncated | 2nd | 0.75 | 0.07 | (-0.09, 0.23) | 0.40 | -0.02 | (-0.20, 0.15) | 0.82 | 0.08 | (-0.07, 0.23) | 0.27 | 167 |
| | 3rd | 0.75 | 0.15 | (-0.03, 0.34) | 0.11 | 0.12 | (-0.07, 0.30) | 0.21 | 0.21 | (0.01, 0.40) | 0.04 | 167 |
| HHV7_U14 | 2nd | 0.50 | 0.00 | (-0.03, 0.03) | 0.97 | 0.01 | (-0.02, 0.04) | 0.44 | 0.01 | (-0.02, 0.04) | 0.65 | 986 |
| | 3rd | 0.50 | -0.01 | (-0.05, 0.02) | 0.51 | -0.01 | (-0.05, 0.02) | 0.51 | -0.01 | (-0.05, 0.02) | 0.38 | 986 |
| HHV7_U14 | 2nd | 0.75 | -0.03 | (-0.10, 0.04) | 0.41 | -0.03 | (-0.07, 0.02) | 0.22 | -0.04 | (-0.11, 0.04) | 0.35 | 986 |
| | 3rd | 0.75 | -0.02 | (-0.09, 0.04) | 0.49 | -0.02 | (-0.08, 0.05) | 0.64 | -0.03 | (-0.10, 0.04) | 0.40 | 986 |
| Tg_p22 | 2nd | 0.50 | 0.01 | (-0.07, 0.08) | 0.87 | 0.00 | (-0.06, 0.07) | 0.90 | 0.02 | (-0.02, 0.06) | 0.37 | 153 |
| | 3rd | 0.50 | 0.07 | (-0.03, 0.18) | 0.17 | 0.04 | (-0.07, 0.15) | 0.52 | 0.09 | (-0.02, 0.19) | 0.10 | 153 |
| Tg_p22 | 2nd | 0.75 | -0.06 | (-0.26, 0.14) | 0.57 | -0.04 | (-0.22, 0.15) | 0.71 | -0.05 | (-0.25, 0.15) | 0.61 | 153 |
| | 3rd | 0.75 | 0.11 | (-0.07, 0.28) | 0.22 | 0.07 | (-0.08, 0.23) | 0.36 | 0.15 | (0.00, 0.30) | 0.05 | 153 |

**eTable 7. Continued.**

| | | | Model 1 | | | Model 2 | | | Model 3 | | | |
|---|---|---|---|---|---|---|---|---|---|---|---|---|
| **Pathogen** | **Tertiles** | **Quantile** | **Coef** | **95% CI** | **p-value** | **Coef** | **95% CI** | **p-value** | **Coef** | **95% CI** | **p-value** | **N** |
| Tg_sag1 | 2nd | 0.50 | 0.02 | (-0.06, 0.10) | 0.56 | 0.03 | (-0.03, 0.09) | 0.35 | 0.03 | (-0.06, 0.11) | 0.53 | 249 |
| | 3rd | 0.50 | 0.04 | (-0.05, 0.12) | 0.40 | 0.05 | (-0.01, 0.11) | 0.10 | 0.04 | (-0.05, 0.12) | 0.39 | 249 |
| Tg_sag1 | 2nd | 0.75 | -0.01 | (-0.15, 0.12) | 0.83 | -0.04 | (-0.13, 0.05) | 0.40 | -0.02 | (-0.15, 0.11) | 0.77 | 249 |
| | 3rd | 0.75 | 0.00 | (-0.11, 0.12) | 0.98 | -0.03 | (-0.15, 0.10) | 0.69 | 0.02 | (-0.09, 0.14) | 0.67 | 249 |
| CpGP3DCT | 2nd | 0.50 | -0.03 | (-0.10, 0.03) | 0.35 | -0.07 | (-0.13, 0.00) | 0.04 | -0.02 | (-0.08, 0.04) | 0.58 | 238 |
| | 3rd | 0.50 | -0.03 | (-0.10, 0.04) | 0.45 | -0.02 | (-0.09, 0.04) | 0.49 | 0.00 | (-0.06, 0.06) | 0.92 | 238 |
| CpGP3DCT | 2nd | 0.75 | -0.07 | (-0.21, 0.07) | 0.32 | -0.09 | (-0.21, 0.03) | 0.15 | -0.05 | (-0.18, 0.07) | 0.40 | 238 |
| | 3rd | 0.75 | -0.02 | (-0.16, 0.13) | 0.82 | 0.03 | (-0.11, 0.17) | 0.71 | -0.04 | (-0.21, 0.12) | 0.62 | 238 |
| HP0547_1 | 2nd | 0.50 | -0.04 | (-0.10, 0.02) | 0.22 | -0.07 | (-0.13, -0.01) | 0.02 | -0.04 | (-0.10, 0.02) | 0.20 | 235 |
| | 3rd | 0.50 | -0.01 | (-0.07, 0.05) | 0.75 | -0.02 | (-0.09, 0.06) | 0.62 | 0.01 | (-0.05, 0.06) | 0.86 | 235 |
| HP0547_1 | 2nd | 0.75 | -0.02 | (-0.18, 0.13) | 0.75 | -0.08 | (-0.19, 0.04) | 0.18 | -0.02 | (-0.20, 0.16) | 0.81 | 235 |
| | 3rd | 0.75 | -0.08 | (-0.23, 0.08) | 0.34 | -0.02 | (-0.12, 0.08) | 0.70 | -0.08 | (-0.26, 0.09) | 0.35 | 235 |
| HP0010 | 2nd | 0.50 | 0.02 | (-0.03, 0.08) | 0.43 | 0.03 | (-0.04, 0.09) | 0.44 | 0.02 | (-0.04, 0.07) | 0.53 | 318 |
| | 3rd | 0.50 | 0.03 | (-0.04, 0.09) | 0.39 | 0.04 | (-0.02, 0.11) | 0.17 | 0.01 | (-0.04, 0.07) | 0.65 | 318 |
| HP0010 | 2nd | 0.75 | 0.07 | (-0.10, 0.25) | 0.41 | -0.02 | (-0.16, 0.12) | 0.74 | 0.04 | (-0.09, 0.17) | 0.51 | 318 |
| | 3rd | 0.75 | 0.04 | (-0.14, 0.21) | 0.68 | -0.04 | (-0.19, 0.10) | 0.57 | 0.01 | (-0.16, 0.18) | 0.91 | 318 |
| HP0073 | 2nd | 0.50 | -0.04 | (-0.10, 0.01) | 0.12 | -0.04 | (-0.11, 0.04) | 0.37 | -0.02 | (-0.09, 0.04) | 0.48 | 210 |
| | 3rd | 0.50 | 0.03 | (-0.08, 0.13) | 0.64 | 0.03 | (-0.05, 0.12) | 0.46 | 0.02 | (-0.08, 0.12) | 0.70 | 210 |
| HP0073 | 2nd | 0.75 | 0.02 | (-0.14, 0.17) | 0.84 | -0.03 | (-0.18, 0.13) | 0.73 | 0.06 | (-0.11, 0.22) | 0.49 | 210 |
| | 3rd | 0.75 | 0.14 | (-0.04, 0.31) | 0.12 | 0.02 | (-0.17, 0.20) | 0.86 | 0.10 | (-0.04, 0.24) | 0.16 | 210 |
| HP0305 | 2nd | 0.50 | -0.04 | (-0.13, 0.04) | 0.30 | -0.08 | (-0.17, 0.01) | 0.09 | -0.01 | (-0.08, 0.07) | 0.84 | 115 |
| | 3rd | 0.50 | 0.07 | (-0.07, 0.21) | 0.30 | 0.07 | (-0.04, 0.18) | 0.23 | 0.08 | (-0.04, 0.21) | 0.20 | 115 |
| HP0305 | 2nd | 0.75 | -0.15 | (-0.30, -0.01) | 0.04 | -0.12 | (-0.29, 0.06) | 0.20 | -0.13 | (-0.27, 0.01) | 0.07 | 115 |
| | 3rd | 0.75 | 0.03 | (-0.10, 0.16) | 0.65 | 0.02 | (-0.09, 0.14) | 0.67 | 0.03 | (-0.09, 0.15) | 0.63 | 115 |
| HP0875 | 2nd | 0.50 | 0.08 | (0.00, 0.17) | 0.05 | 0.07 | (-0.02, 0.15) | 0.15 | 0.06 | (-0.01, 0.13) | 0.07 | 171 |
| | 3rd | 0.50 | 0.06 | (-0.05, 0.17) | 0.28 | 0.03 | (-0.07, 0.12) | 0.59 | 0.05 | (-0.07, 0.17) | 0.40 | 171 |
| HP0875 | 2nd | 0.75 | 0.02 | (-0.21, 0.25) | 0.87 | 0.20 | (-0.02, 0.43) | 0.07 | 0.00 | (-0.26, 0.27) | 0.98 | 171 |
| | 3rd | 0.75 | 0.05 | (-0.15, 0.25) | 0.62 | 0.10 | (-0.09, 0.28) | 0.29 | 0.10 | (-0.16, 0.35) | 0.44 | 171 |
| HP0887_2 | 2nd | 0.50 | -0.10 | (-0.20, 0.01) | 0.08 | -0.09 | (-0.17, 0.00) | 0.04 | -0.09 | (-0.17, -0.01) | 0.02 | 126 |
| | 3rd | 0.50 | -0.07 | (-0.20, 0.06) | 0.31 | -0.02 | (-0.11, 0.07) | 0.67 | -0.06 | (-0.18, 0.06) | 0.34 | 126 |
| HP0887_2 | 2nd | 0.75 | -0.15 | (-0.34, 0.04) | 0.13 | -0.11 | (-0.35, 0.13) | 0.36 | -0.15 | (-0.37, 0.07) | 0.19 | 126 |
| | 3rd | 0.75 | 0.05 | (-0.20, 0.29) | 0.71 | 0.04 | (-0.22, 0.31) | 0.74 | 0.05 | (-0.21, 0.30) | 0.72 | 126 |

**eTable 7. Continued.**

| | | | Model 1 | | | Model 2 | | | Model 3 | | | |
|---|---|---|---|---|---|---|---|---|---|---|---|---|
| **Pathogen** | **Tertiles** | **Quantile** | **Coef** | **95% CI** | **p-value** | **Coef** | **95% CI** | **p-value** | **Coef** | **95% CI** | **p-value** | **N** |
| | 3rd | 0.50 | 0.05 | (-0.08, 0.17) | 0.48 | 0.05 | (-0.03, 0.13) | 0.21 | 0.04 | (-0.08, 0.15) | 0.53 | 132 |
| HP1098 | 2nd | 0.75 | -0.11 | (-0.28, 0.06) | 0.19 | 0.00 | (-0.15, 0.15) | 0.99 | -0.05 | (-0.24, 0.13) | 0.57 | 132 |
| | 3rd | 0.75 | -0.09 | (-0.29, 0.12) | 0.41 | 0.05 | (-0.13, 0.23) | 0.62 | -0.06 | (-0.28, 0.17) | 0.62 | 132 |
| HP1564 | 2nd | 0.50 | -0.01 | (-0.08, 0.05) | 0.66 | 0.00 | (-0.07, 0.06) | 0.93 | -0.01 | (-0.08, 0.05) | 0.68 | 260 |
| | 3rd | 0.50 | -0.02 | (-0.11, 0.06) | 0.55 | -0.04 | (-0.10, 0.03) | 0.26 | -0.05 | (-0.13, 0.03) | 0.24 | 260 |
| HP1564 | 2nd | 0.75 | 0.00 | (-0.15, 0.15) | 0.97 | 0.03 | (-0.13, 0.19) | 0.70 | 0.04 | (-0.10, 0.17) | 0.60 | 260 |
| | 3rd | 0.75 | 0.00 | (-0.17, 0.16) | 0.95 | 0.00 | (-0.17, 0.17) | 0.99 | 0.05 | (-0.10, 0.19) | 0.54 | 260 |
| BK_VP1 | 2nd | 0.50 | -0.01 | (-0.03, 0.02) | 0.59 | 0.00 | (-0.03, 0.03) | 0.93 | 0.00 | (-0.03, 0.03) | 0.97 | 1244 |
| | 3rd | 0.50 | -0.02 | (-0.05, 0.01) | 0.18 | -0.01 | (-0.04, 0.02) | 0.53 | -0.02 | (-0.05, 0.01) | 0.23 | 1244 |
| BK_VP1 | 2nd | 0.75 | 0.02 | (-0.05, 0.10) | 0.55 | 0.01 | (-0.05, 0.06) | 0.81 | 0.01 | (-0.06, 0.09) | 0.71 | 1244 |
| | 3rd | 0.75 | -0.02 | (-0.08, 0.03) | 0.42 | 0.00 | (-0.05, 0.05) | 0.94 | -0.02 | (-0.08, 0.04) | 0.51 | 1244 |
| JC_VP1 | 2nd | 0.50 | 0.04 | (0.01, 0.08) | 0.01 | 0.04 | (0.00, 0.09) | 0.03 | 0.04 | (0.01, 0.07) | 0.02 | 706 |
| | 3rd | 0.50 | 0.01 | (-0.02, 0.05) | 0.42 | 0.01 | (-0.03, 0.04) | 0.70 | 0.01 | (-0.02, 0.05) | 0.37 | 706 |
| JC_VP1 | 2nd | 0.75 | 0.06 | (-0.01, 0.14) | 0.12 | 0.05 | (0.00, 0.10) | 0.05 | 0.06 | (-0.02, 0.13) | 0.12 | 706 |
| | 3rd | 0.75 | 0.05 | (-0.03, 0.13) | 0.22 | 0.05 | (-0.03, 0.12) | 0.21 | 0.07 | (-0.02, 0.16) | 0.11 | 706 |
| MCV344_VP1 | 2nd | 0.50 | 0.02 | (-0.02, 0.05) | 0.34 | -0.01 | (-0.04, 0.02) | 0.54 | 0.01 | (-0.02, 0.04) | 0.50 | 812 |
| | 3rd | 0.50 | 0.00 | (-0.03, 0.03) | 0.91 | -0.02 | (-0.05, 0.02) | 0.30 | 0.00 | (-0.03, 0.03) | 1.00 | 812 |
| MCV344_VP1 | 2nd | 0.75 | 0.01 | (-0.04, 0.06) | 0.68 | 0.02 | (-0.03, 0.07) | 0.51 | 0.01 | (-0.05, 0.07) | 0.71 | 812 |
| | 3rd | 0.75 | 0.02 | (-0.04, 0.07) | 0.56 | 0.02 | (-0.05, 0.09) | 0.60 | 0.02 | (-0.05, 0.09) | 0.58 | 812 |

Note: quantile regression results depicting differences in p-tau217 concentrations at the 50th and 75th quantiles of the p-tau distribution per unit of exposure -- seropositive with reference to seronegative for individual pathogens, or per 1 additional pathogen in the pathogen burden indices. Model 1 adjusted for sex, age at serology, and age at p-tau217 measurement. Model 2 adjusts for model 1 covariates plus APOE ε4 carriage. Model 3 adjusts for model 1 covariates plus education.

Abbreviations: HSV1 = Herpes simplex virus-1; HSV2 = Herpes simplex virus-2; VZV = Varicella zoster virus; EBV = Epstein-Barr virus; CMV = Cytomegalovirus; HHV6A = Human herpesvirus-6a; HHV6B = Human herpesvirus-6b; HHV7 = Human herpesvirus-7; BK = BK virus; JC = JC virus; MCV = Merkel cell polyomavirus; Tg = Toxoplasma gondii; Hp = Helicobacter pylori; Ct = Chlamydia trachomatis; PBI = Pathogen burden Index.

**eTable 8. Interaction terms from tests of interactions of HSV1 serostatus with both VZV and CMV serostatus in relation to median p-tau217.**

| | Model 1 | | | Model 2 | | | Model 3 | | |
|---|---|---|---|---|---|---|---|---|---|
| | **Coeff** | **95% CI** | **p-value** | **Coeff** | **95% CI** | **p-value** | **Coeff** | **95% CI** | **p-value** |
| HSV1xVZV serostatus | -0.036 | (-0.10, 0.03) | 0.268 | -0.022 | (-0.08, 0.04) | 0.475 | -0.041 | (-0.11, 0.03) | 0.248 |
| HSV1xCMV serostatus | -0.012 | (-0.06, 0.04) | 0.630 | 0.008 | (-0.04, 0.06) | 0.770 | -0.023 | (-0.07, 0.02) | 0.300 |

Note: Model 1 adjusted for sex, age at serology, and age at p-tau217 measurement. Model 2 adjusted for model 1 covariates plus APOE ε4 carriage. Model 3 adjusted for model 1 covariates plus education.

Abbreviations: HSV1 = Herpes simplex virus-1; VZV = Varicella zoster virus; EBV = Epstein-Barr virus; CMV = Cytomegalovirus

**eTable 9. Interaction terms from tests of interactions of pathogen serostatus and pathogen burden with APOE ε4 carriage and educational attainment in relation to median p-tau217 concentration**

| | Interaction terms for exposure x *APOE* ε4 carriage | | | Interaction terms for exposure x education | | |
|---|---|---|---|---|---|---|
| | **Coeff** | **95% CI** | **p-value** | **Coeff** | **95% CI** | **p-value** |
| **Pathogen serostatus** | | | | | | |
| HSV1 | 0.014 | (-0.05, 0.08) | 0.674 | -0.023 | (-0.07, 0.02) | 0.293 |
| HSV2 | 0.003 | (-0.06, 0.06) | 0.911 | -0.007 | (-0.08, 0.07) | 0.863 |
| VZV | 0.010 | (-0.08, 0.10) | 0.839 | -0.014 | (-0.07, 0.04) | 0.634 |
| EBV | 0.095 | (0.01, 0.18) | **0.024** | -0.032 | (-0.10, 0.04) | 0.387 |
| CMV | 0.063 | (0.00, 0.13) | **0.055** | -0.007 | (-0.05, 0.04) | 0.755 |
| HHV6A | -0.024 | (-0.08, 0.04) | 0.427 | 0.033 | (-0.01, 0.08) | 0.152 |
| HHV6B | 0.055 | (-0.01, 0.12) | **0.090** | -0.002 | (-0.04, 0.04) | 0.935 |
| HHV7 | 0.036 | (-0.04, 0.12) | 0.380 | -0.039 | (-0.09, 0.01) | 0.115 |
| BK | 0.008 | (-0.22, 0.24) | 0.948 | 0.053 | (-0.09, 0.20) | 0.479 |
| JC | -0.012 | (-0.07, 0.05) | 0.705 | 0.027 | (-0.02, 0.07) | 0.237 |
| MCV | 0.007 | (-0.07, 0.08) | 0.853 | 0.004 | (-0.04, 0.05) | 0.851 |
| Tg | 0.046 | (-0.02, 0.12) | 0.191 | 0.008 | (-0.04, 0.06) | 0.769 |
| Hp | 0.089 | (0.00, 0.18) | **0.049** | -0.080 | (-0.13, -0.03) | **0.001** |
| Ct | -0.003 | (-0.07, 0.06) | 0.928 | -0.039 | (-0.09, 0.01) | 0.137 |
| **Pathogen burden** | | | | | | |
| Total | 0.013 | (0.00, 0.03) | **0.102** | -0.003 | (-0.01, 0.01) | 0.590 |
| Neurotropic | 0.011 | (-0.01, 0.03) | 0.240 | -0.003 | (-0.02, 0.01) | 0.638 |

Note: Interaction terms were added to model 1 adjusted for sex, age at serology, and age at p-tau217 measurement.

Abbreviations: HSV1 = Herpes simplex virus-1; HSV2 = Herpes simplex virus-2; VZV = Varicella zoster virus; EBV = Epstein-Barr virus; CMV = Cytomegalovirus; HHV6A = Human herpesvirus-6a; HHV6B = Human herpesvirus-6b; HHV7 = Human herpesvirus-7; BK = BK virus; JC = JC virus; MCV = Merkel cell polyomavirus; Tg = Toxoplasma gondii; Hp = Helicobacter pylori; Ct = Chlamydia trachomatis.

**eTable 10. Interaction terms from tests of interactions of pathogen antigen seroreactivities with *APOE* ε4 carriage and educational attainment in relation to median p-tau217 concentration**

| | | Interaction terms for exposure x *APOE* ε4 carriage | | | | Interaction terms for exposure x education | | | |
|---|---|---|---|---|---|---|---|---|---|
| **Antigen** | **Tertiles** | **Coef** | **95% CI** | **p-value** | **Testparm p-value** | **Coef** | **95% CI** | **p-value** | **Testparm p-value** |
| HSV1_1gG | 2nd | 0.020 | (-0.08, 0.12) | 0.691 | 0.698 | 0.013 | (-0.06, 0.09) | 0.732 | 0.492 |
| | 3rd | -0.020 | (-0.10, 0.06) | 0.628 | | 0.041 | (-0.03, 0.11) | 0.252 | |
| HSV2_2mgGunique | 2nd | 0.040 | (-0.09, 0.17) | 0.535 | 0.824 | -0.129 | (-0.33, 0.07) | 0.211 | 0.106 |
| | 3rd | 0.010 | (-0.10, 0.12) | 0.858 | | 0.082 | (-0.07, 0.23) | 0.276 | |
| VZV_gE_gI | 2nd | -0.118 | (-0.21, -0.02) | 0.013 | **0.046** | 0.008 | (-0.05, 0.07) | 0.801 | 0.514 |
| | 3rd | -0.088 | (-0.18, 0.01) | 0.067 | | 0.033 | (-0.03, 0.09) | 0.278 | |
| EBV_VCAp18 | 2nd | -0.048 | (-0.14, 0.04) | 0.295 | 0.376 | -0.025 | (-0.08, 0.03) | 0.372 | 0.514 |
| | 3rd | 0.001 | (-0.10, 0.10) | 0.976 | | 0.006 | (-0.05, 0.06) | 0.828 | |
| EBV_EBNA_peptide | 2nd | 0.059 | (-0.02, 0.14) | 0.150 | 0.292 | 0.015 | (-0.04, 0.07) | 0.579 | 0.563 |
| | 3rd | 0.003 | (-0.08, 0.09) | 0.947 | | -0.014 | (-0.07, 0.04) | 0.649 | |
| EBV_Zebra | 2nd | -0.006 | (-0.09, 0.08) | 0.888 | 0.741 | 0.038 | (-0.02, 0.09) | 0.170 | **0.053** |
| | 3rd | -0.035 | (-0.13, 0.06) | 0.448 | | 0.063 | (0.01, 0.11) | 0.016 | |
| EBV_EAD | 2nd | -0.033 | (-0.14, 0.08) | 0.549 | 0.659 | 0.033 | (-0.02, 0.09) | 0.232 | 0.442 |
| | 3rd | -0.051 | (-0.16, 0.06) | 0.365 | | 0.033 | (-0.03, 0.10) | 0.311 | |
| CMV_pp150NTerm | 2nd | 0.063 | (-0.10, 0.22) | 0.448 | 0.746 | -0.028 | (-0.11, 0.05) | 0.486 | 0.494 |
| | 3rd | 0.040 | (-0.11, 0.19) | 0.594 | | 0.016 | (-0.05, 0.08) | 0.658 | |
| CMV_pp52 | 2nd | 0.048 | (-0.07, 0.17) | 0.423 | 0.361 | 0.015 | (-0.06, 0.09) | 0.701 | 0.801 |
| | 3rd | 0.091 | (-0.03, 0.22) | 0.156 | | -0.009 | (-0.09, 0.07) | 0.809 | |
| CMV_pp28 | 2nd | 0.095 | (-0.02, 0.21) | 0.113 | **0.080** | 0.003 | (-0.05, 0.06) | 0.914 | 0.418 |
| | 3rd | 0.116 | (-0.01, 0.24) | 0.065 | | -0.040 | (-0.11, 0.03) | 0.282 | |
| HHV6_IE1A_truncated | 2nd | 0.082 | (-0.07, 0.23) | 0.290 | 0.571 | -0.031 | (-0.14, 0.08) | 0.595 | 0.558 |
| | 3rd | 0.063 | (-0.13, 0.25) | 0.510 | | 0.022 | (-0.07, 0.11) | 0.639 | |
| HHV6_p100truncated | 2nd | 0.028 | (-0.35, 0.41) | 0.885 | 0.718 | -0.039 | (-0.31, 0.23) | 0.779 | 0.686 |
| | 3rd | -0.103 | (-0.38, 0.17) | 0.456 | | 0.060 | (-0.10, 0.22) | 0.468 | |
| HHV6_IE1B_truncated | 2nd | 0.043 | (-0.07, 0.16) | 0.460 | 0.335 | -0.035 | (-0.12, 0.05) | 0.430 | 0.629 |
| | 3rd | 0.076 | (-0.03, 0.18) | 0.146 | | 0.010 | (-0.08, 0.10) | 0.828 | |
| HHV6_p101Ktruncated | 2nd | 0.165 | (-0.01, 0.34) | 0.070 | 0.170 | 0.084 | (-0.07, 0.24) | 0.282 | 0.533 |
| | 3rd | 0.095 | (-0.14, 0.33) | 0.428 | | 0.037 | (-0.11, 0.19) | 0.631 | |
| HHV7_U14 | 2nd | 0.016 | (-0.08, 0.11) | 0.745 | 0.804 | 0.009 | (-0.05, 0.07) | 0.759 | 0.855 |
| | 3rd | -0.013 | (-0.10, 0.08) | 0.771 | | -0.006 | (-0.07, 0.06) | 0.859 | |

**eTable 10. Continued**

| Antigen | Tertiles | Interaction terms for exposure x *APOE* ε4 carriage | | | | Interaction terms for exposure x education | | | |
|---|---|---|---|---|---|---|---|---|---|
| | | **Coef** | **95% CI** | **p-value** | **Testparm p-value** | **Coef** | **95% CI** | **p-value** | **Testparm p-value** |
| Tg_p22 | 2nd | 0.053 | (-0.09, 0.20) | 0.474 | 0.729 | -0.055 | (-0.13, 0.02) | 0.147 | 0.294 |
| | 3rd | 0.082 | (-0.16, 0.32) | 0.499 | | 0.028 | (-0.16, 0.22) | 0.773 | |
| Tg_sag1 | 2nd | 0.034 | (-0.16, 0.22) | 0.728 | 0.885 | 0.008 | (-0.15, 0.16) | 0.922 | 0.970 |
| | 3rd | -0.005 | (-0.17, 0.15) | 0.948 | | -0.010 | (-0.17, 0.15) | 0.902 | |
| Ct_pGP3DCT | 2nd | 0.024 | (-0.11, 0.16) | 0.727 | 0.876 | 0.078 | (-0.04, 0.20) | 0.196 | 0.376 |
| | 3rd | 0.042 | (-0.12, 0.21) | 0.610 | | 0.063 | (-0.04, 0.17) | 0.251 | |
| HP0547_1 | 2nd | 0.007 | (-0.21, 0.23) | 0.948 | 0.508 | -0.041 | (-0.19, 0.11) | 0.590 | 0.594 |
| | 3rd | -0.114 | (-0.37, 0.14) | 0.380 | | -0.072 | (-0.21, 0.07) | 0.308 | |
| HP0010 | 2nd | 0.049 | (-0.13, 0.23) | 0.592 | 0.858 | -0.065 | (-0.19, 0.06) | 0.294 | 0.425 |
| | 3rd | 0.036 | (-0.14, 0.21) | 0.681 | | -0.074 | (-0.19, 0.05) | 0.229 | |
| HP0073 | 2nd | 0.065 | (-0.43, 0.56) | 0.796 | 0.768 | 0.031 | (-0.09, 0.15) | 0.624 | **0.034** |
| | 3rd | 0.133 | (-0.33, 0.60) | 0.575 | | -0.194 | (-0.36, -0.03) | 0.023 | |
| HP0305 | 2nd | -0.016 | (-0.25, 0.22) | 0.894 | 0.299 | 0.086 | (-0.09, 0.26) | 0.340 | 0.367 |
| | 3rd | 0.076 | (-0.17, 0.32) | 0.540 | | 0.200 | (-0.09, 0.49) | 0.177 | |
| HP0875 | 2nd | -0.179 | (-0.39, 0.03) | 0.099 | 0.186 | -0.098 | (-0.29, 0.10) | 0.320 | 0.476 |
| | 3rd | -0.004 | (-0.28, 0.27) | 0.975 | | 0.018 | (-0.23, 0.27) | 0.889 | |
| HP0887_2 | 2nd | 0.003 | (-0.22, 0.22) | 0.977 | 0.265 | 0.009 | (-0.18, 0.20) | 0.925 | 0.989 |
| | 3rd | 0.189 | (-0.05, 0.43) | 0.124 | | 0.015 | (-0.19, 0.22) | 0.886 | |
| HP1098 | 2nd | -0.084 | (-0.28, 0.11) | 0.388 | **0.040** | 0.340 | (0.05, 0.63) | 0.022 | **0.072** |
| | 3rd | -0.216 | (-0.40, -0.03) | 0.023 | | 0.191 | (-0.09, 0.47) | 0.181 | |
| HP1564 | 2nd | 0.040 | (-0.18, 0.26) | 0.725 | 0.697 | 0.093 | (-0.03, 0.22) | 0.148 | 0.348 |
| | 3rd | -0.059 | (-0.22, 0.10) | 0.476 | | 0.011 | (-0.15, 0.17) | 0.893 | |
| BK_VP1 | 2nd | 0.003 | (-0.08, 0.09) | 0.936 | 0.184 | 0.037 | (-0.01, 0.09) | 0.144 | 0.336 |
| | 3rd | 0.061 | (-0.01, 0.13) | 0.092 | | 0.009 | (-0.05, 0.06) | 0.750 | |
| JC_VP1 | 2nd | 0.027 | (-0.07, 0.13) | 0.592 | 0.818 | 0.042 | (-0.03, 0.11) | 0.252 | 0.518 |
| | 3rd | -0.004 | (-0.09, 0.08) | 0.923 | | 0.011 | (-0.06, 0.08) | 0.740 | |
| MCV344_VP1 | 2nd | 0.017 | (-0.08, 0.11) | 0.729 | 0.883 | -0.032 | (-0.09, 0.03) | 0.281 | **0.028** |
| | 3rd | -0.008 | (-0.11, 0.09) | 0.867 | | 0.057 | (-0.02, 0.13) | 0.126 | |

Note: Interaction terms were added to model 1 adjusted for sex, age at serology, and age at p-tau217 measurement. Abbreviations: HSV1 = Herpes simplex virus-1; HSV2 = Herpes simplex virus-2; VZV = Varicella zoster virus; EBV = Epstein-Barr virus; CMV = Cytomegalovirus;

HHV6A = Human herpesvirus-6a; HHV6B = Human herpesvirus-6b; HHV7 = Human herpesvirus-7; BK = BK virus; JC = JC virus; MCV = Merkel cell polyomavirus; Tg = Toxoplasma gondii; Hp = Helicobacter pylori; Ct = Chlamydia trachomatis.

**eTable 11. Associations of pathogen serostatus, pathogen burden and seroreactivities with median p-tau217 stratified by APOE ε4 carriage. Results are presented for Model 1, adjusted for age at serology, age at p-tau217 measurement, and sex.**

| | | non-ε4 carriers | | | | ε4 carriers | | | |
|---|---|---|---|---|---|---|---|---|---|
| | | **Coef** | **95% CI** | **p-value** | **N** | **Coef** | **95% CI** | **p-value** | **N** |
| **Pathogen serostatus** | | | | | | | | | |
| EBV | | -0.039 | (-0.11, 0.03) | 0.300 | 945 | 0.037 | (-0.03, 0.10) | 0.249 | 411 |
| CMV | | -0.009 | (-0.03, 0.01) | 0.467 | 945 | 0.077 | (0.01, 0.14) | 0.017 | 411 |
| HHV6B | | -0.019 | (-0.04, 0.01) | 0.138 | 945 | 0.042 | (-0.02, 0.11) | 0.208 | 411 |
| Hp | | 0.003 | (-0.03, 0.04) | 0.833 | 945 | 0.084 | (0.01, 0.16) | 0.035 | 411 |
| **Pathogen burden** | | | | | | | | | |
| Total | | -0.007 | (-0.01, 0.00) | 0.033 | 945 | 0.010 | (0.00, 0.02) | 0.175 | 411 |
| **Seroreactivity** | **Tertiles** | | | | | | | | |
| VZV_gE_gI | 2nd | 0.014 | (-0.02, 0.05) | 0.431 | | -0.123 | (-0.21, -0.03) | 0.008 | |
| | 3rd | 0.002 | (-0.03, 0.03) | 0.920 | 740 | -0.105 | (-0.20, -0.02) | 0.021 | 337 |
| CMV_pp28 | 2nd | 0.003 | (-0.03, 0.04) | 0.856 | | 0.093 | (-0.04, 0.22) | 0.155 | |
| | 3rd | 0.011 | (-0.03, 0.05) | 0.578 | 533 | 0.132 | (0.02, 0.24) | 0.020 | 233 |
| HP1098 | 2nd | 0.010 | (-0.05, 0.07) | 0.761 | | -0.003 | (-0.17, 0.17) | 0.969 | |
| | 3rd | 0.069 | (0.00, 0.13) | 0.038 | 91 | -0.058 | (-0.19, 0.08) | 0.391 | 41 |

Note: Results are presented for Model 1, adjusted for age at serology, age at p-tau217 measurement, and sex.

Abbreviations: VZV = Varicella zoster virus; EBV = Epstein-Barr virus; CMV = Cytomegalovirus; HHV6B = Human herpesvirus-6b; Hp = Helicobacter pylori.

**eTable 12. Associations of pathogen serostatus and seroreactivities with median p-tau217 stratified by educational attainment. Results are presented for Model 1, adjusted for age at serology, age at p-tau217 measurement, and sex.**

| | | O-levels or equivalent or below | | | | A-levels or higher | | | |
|---|---|---|---|---|---|---|---|---|---|
| | | **Coef** | **95% CI** | **p-value** | **N** | **Coef** | **95% CI** | **p-value** | **N** |
| **Pathogen serostatus** | | | | | | | | | |
| Hp | | 0.040 | (0.01, 0.07) | 0.021 | 698 | -0.050 | (-0.09, -0.01) | 0.005 | 658 |
| **Seroreactivity** | **Tertiles** | | | | | | | | |
| EBV_Zebra | 2nd | -0.008 | (-0.05, 0.03) | 0.675 | | 0.024 | (-0.01, 0.06) | 0.215 | |
| | 3rd | -0.023 | (-0.06, 0.01) | 0.182 | 638 | 0.033 | (-0.01, 0.07) | 0.109 | 563 |
| HP0073 | 2nd | -0.018 | (-0.10, 0.06) | 0.654 | | -0.021 | (-0.14, 0.10) | 0.721 | |
| | 3rd | 0.125 | (0.00, 0.25) | 0.051 | 106 | -0.074 | (-0.19, 0.05) | 0.225 | 104 |
| HP1098 | 2nd | -0.192 | (-0.40, 0.01) | 0.064 | | 0.152 | (0.04, 0.27) | 0.011 | |
| | 3rd | -0.127 | (-0.33, 0.08) | 0.221 | 76 | 0.097 | (-0.07, 0.27) | 0.252 | 56 |
| MCV344_VP1 | 2nd | 0.014 | (-0.04, 0.06) | 0.602 | | -0.007 | (-0.05, 0.04) | 0.774 | |
| | 3rd | -0.036 | (-0.10, 0.02) | 0.246 | 406 | 0.028 | (-0.02, 0.07) | 0.242 | 406 |

Note: Results are presented for Model 1, adjusted for age at serology, age at p-tau217 measurement, and sex.

Abbreviations: EBV = Epstein-Barr virus; MCV = Merkel cell polyomavirus; Hp = Helicobacter pylori.

**eTable 13. Associations of pathogen serostatus and pathogen burden with odds of cerebral amyloidosis measured by Aβ-PET.**

| | Model 1 | | | Model 2 | | | Model 3 | | |
|---|---|---|---|---|---|---|---|---|---|
| **Exposure** | **ORs** | **95% CI** | **p-value** | **ORs** | **95% CI** | **p-value** | **ORs** | **95% CI** | **p-value** |
| HSV1 | 1.05 | (0.60, 1.83) | 0.86 | 1.21 | (0.68, 2.17) | 0.52 | 1.06 | (0.61, 1.84) | 0.85 |
| HSV2 | 0.72 | (0.24, 2.16) | 0.56 | 0.63 | (0.16, 2.39) | 0.49 | 0.72 | (0.24, 2.17) | 0.56 |
| VZV | 1.36 | (0.71, 2.60) | 0.35 | 1.12 | (0.56, 2.25) | 0.75 | 1.38 | (0.72, 2.63) | 0.33 |
| EBV | 2.84 | (0.95, 8.52) | 0.06 | 3.11 | (0.96, 10.09) | 0.06 | 2.91 | (0.97, 8.72) | 0.06 |
| CMV | 0.92 | (0.55, 1.53) | 0.75 | 0.93 | (0.54, 1.61) | 0.80 | 0.93 | (0.55, 1.55) | 0.77 |
| HHV6A | 0.96 | (0.57, 1.63) | 0.88 | 0.99 | (0.56, 1.75) | 0.98 | 0.96 | (0.57, 1.63) | 0.88 |
| HHV6B | 0.94 | (0.56, 1.58) | 0.82 | 0.88 | (0.51, 1.52) | 0.65 | 0.93 | (0.56, 1.57) | 0.80 |
| HHV7 | 0.76 | (0.43, 1.35) | 0.35 | 0.84 | (0.45, 1.58) | 0.59 | 0.76 | (0.43, 1.34) | 0.34 |
| BK | 0.92 | (0.36, 2.36) | 0.86 | 0.99 | (0.37, 2.70) | 0.99 | 0.91 | (0.35, 2.33) | 0.84 |
| JC | 1.08 | (0.65, 1.81) | 0.77 | 1.12 | (0.65, 1.95) | 0.68 | 1.08 | (0.65, 1.81) | 0.77 |
| MCV | 0.83 | (0.48, 1.42) | 0.50 | 0.73 | (0.41, 1.29) | 0.28 | 0.83 | (0.48, 1.42) | 0.49 |
| Tg | 1.69 | (0.94, 3.05) | 0.08 | 1.60 | (0.87, 2.94) | 0.13 | 1.69 | (0.94, 3.05) | 0.08 |
| Hp | 1.15 | (0.57, 2.31) | 0.69 | 1.16 | (0.56, 2.41) | 0.69 | 1.16 | (0.58, 2.32) | 0.68 |
| Ct | 0.85 | (0.39, 1.86) | 0.69 | 0.85 | (0.38, 1.92) | 0.70 | 0.86 | (0.39, 1.88) | 0.70 |
| Total PB | 1.01 | (0.88, 1.17) | 0.85 | 1.01 | (0.86, 1.18) | 0.90 | 1.01 | (0.88, 1.17) | 0.84 |
| Neurotropic PB | 1.03 | (0.88, 1.22) | 0.68 | 1.04 | (0.87, 1.24) | 0.68 | 1.04 | (0.88, 1.22) | 0.66 |

Note: Model 1 adjusted for age at serology, age at scan, and sex; Model 2 adjusted for model 1 covariates plus APOE ε4 carriage; Model 3 adjusted for model 1 covariates plus Education. These results are depicted in eFigure 3

Abbreviations: HSV1 = Herpes simplex virus-1; HSV2 = Herpes simplex virus-2; VZV = Varicella zoster virus; EBV = Epstein-Barr virus; CMV = Cytomegalovirus; HHV6A = Human herpesvirus-6a; HHV6B = Human herpesvirus-6b; HHV7 = Human herpesvirus-7; BK = BK virus; JC = JC virus; MCV = Merkel cell polyomavirus; Tg = Toxoplasma gondii; Hp = Helicobacter pylori; Ct = Chlamydia trachomatis; PBI = Pathogen burden index.

**eTable 14: Associations of tertiles of pathogen antigen seroreactivities with odds of cerebral amyloidosis measured by Aβ-PET**

| | | Model 1 | | | Model 2 | | | Model 3 | | |
|---|---|---|---|---|---|---|---|---|---|---|
| **Antigens** | **Tertiles** | **ORs** | **95% CI** | **p-value** | **ORs** | **95% CI** | **p-value** | **ORs** | **95% CI** | **p-value** |
| HSV1_1gG | 2nd | 0.87 | (0.41, 1.85) | 0.72 | 0.97 | (0.45, 2.11) | 0.94 | 0.88 | (0.41, 1.86) | 0.73 |
| HSV1_1gG | 3rd | 0.89 | (0.42, 1.91) | 0.77 | 1.10 | (0.48, 2.51) | 0.83 | 0.89 | (0.42, 1.91) | 0.77 |
| VZV_gE_gI | 2nd | 0.86 | (0.43, 1.70) | 0.66 | 0.80 | (0.38, 1.67) | 0.55 | 0.85 | (0.43, 1.69) | 0.65 |
| VZV_gE_gI | 3rd | 0.62 | (0.31, 1.27) | 0.19 | 0.58 | (0.28, 1.22) | 0.15 | 0.62 | (0.30, 1.26) | 0.18 |
| EBV_VCAp18 | 2nd | 1.54 | (0.79, 3.00) | 0.21 | 1.70 | (0.81, 3.53) | 0.16 | 1.53 | (0.78, 3.00) | 0.21 |
| EBV_VCAp18 | 3rd | 1.45 | (0.74, 2.83) | 0.27 | 1.76 | (0.91, 3.43) | 0.10 | 1.44 | (0.74, 2.83) | 0.29 |
| EBV_EBNA_peptide | 2nd | 0.46 | (0.23, 0.92) | 0.03 | 0.49 | (0.24, 1.02) | 0.06 | 0.46 | (0.23, 0.92) | 0.03 |
| EBV_EBNA_peptide | 3rd | 0.72 | (0.38, 1.37) | 0.32 | 0.71 | (0.36, 1.41) | 0.33 | 0.72 | (0.38, 1.37) | 0.32 |
| EBV_Zebra | 2nd | 0.96 | (0.48, 1.93) | 0.92 | 0.85 | (0.40, 1.80) | 0.68 | 0.96 | (0.48, 1.92) | 0.91 |
| EBV_Zebra | 3rd | 1.13 | (0.57, 2.23) | 0.73 | 1.28 | (0.60, 2.71) | 0.52 | 1.13 | (0.57, 2.23) | 0.72 |
| EBV_EAD | 2nd | 1.51 | (0.78, 2.93) | 0.23 | 1.58 | (0.78, 3.22) | 0.20 | 1.50 | (0.77, 2.92) | 0.23 |
| EBV_EAD | 3rd | 1.04 | (0.52, 2.08) | 0.91 | 1.07 | (0.51, 2.22) | 0.86 | 1.03 | (0.51, 2.06) | 0.94 |
| CMV_pp150NTerm | 2nd | 1.12 | (0.48, 2.60) | 0.79 | 0.74 | (0.30, 1.86) | 0.53 | 1.12 | (0.48, 2.60) | 0.80 |
| CMV_pp150NTerm | 3rd | 1.34 | (0.57, 3.12) | 0.50 | 1.21 | (0.47, 3.14) | 0.69 | 1.33 | (0.57, 3.13) | 0.51 |
| CMV_pp52 | 2nd | 0.47 | (0.19, 1.17) | 0.11 | 0.56 | (0.20, 1.55) | 0.26 | 0.46 | (0.18, 1.17) | 0.10 |
| CMV_pp52 | 3rd | 0.81 | (0.35, 1.90) | 0.63 | 0.85 | (0.35, 2.03) | 0.71 | 0.80 | (0.34, 1.87) | 0.60 |
| CMV_pp28 | 2nd | 0.88 | (0.38, 2.02) | 0.76 | 0.86 | (0.35, 2.11) | 0.75 | 0.89 | (0.38, 2.07) | 0.79 |
| CMV_pp28 | 3rd | 0.83 | (0.36, 1.91) | 0.66 | 0.73 | (0.30, 1.76) | 0.48 | 0.82 | (0.35, 1.92) | 0.65 |
| HHV6_IE1A_truncated | 2nd | 1.48 | (0.55, 4.00) | 0.44 | 1.14 | (0.40, 3.28) | 0.80 | 1.45 | (0.54, 3.91) | 0.46 |
| HHV6_IE1A_truncated | 3rd | 0.84 | (0.29, 2.45) | 0.76 | 0.82 | (0.27, 2.51) | 0.73 | 0.87 | (0.30, 2.52) | 0.80 |
| HHV6_IE1B_truncated | 2nd | 0.86 | (0.33, 2.22) | 0.76 | 0.93 | (0.33, 2.58) | 0.88 | 0.86 | (0.33, 2.23) | 0.76 |
| HHV6_IE1B_truncated | 3rd | 1.35 | (0.54, 3.40) | 0.52 | 2.18 | (0.86, 5.56) | 0.10 | 1.35 | (0.54, 3.39) | 0.53 |
| HHV7_U14 | 2nd | 1.71 | (0.79, 3.69) | 0.17 | 1.99 | (0.88, 4.48) | 0.10 | 1.69 | (0.78, 3.67) | 0.18 |
| HHV7_U14 | 3rd | 1.32 | (0.61, 2.87) | 0.48 | 1.27 | (0.52, 3.08) | 0.60 | 1.29 | (0.59, 2.81) | 0.52 |
| BK_VP1 | 2nd | 1.31 | (0.67, 2.55) | 0.43 | 1.43 | (0.71, 2.87) | 0.31 | 1.31 | (0.67, 2.54) | 0.43 |
| BK_VP1 | 3rd | 1.38 | (0.72, 2.62) | 0.33 | 1.25 | (0.61, 2.57) | 0.54 | 1.36 | (0.72, 2.59) | 0.35 |
| JC_VP1 | 2nd | 0.82 | (0.36, 1.91) | 0.65 | 0.83 | (0.36, 1.93) | 0.66 | 0.82 | (0.36, 1.90) | 0.65 |
| JC_VP1 | 3rd | 0.64 | (0.26, 1.54) | 0.32 | 0.67 | (0.26, 1.71) | 0.40 | 0.63 | (0.26, 1.54) | 0.32 |
| MCV344_VP | 2nd | 1.17 | (0.52, 2.64) | 0.70 | 0.94 | (0.39, 2.25) | 0.89 | 1.18 | (0.52, 2.66) | 0.69 |
| MCV344_VP | 3rd | 1.81 | (0.83, 3.95) | 0.14 | 1.53 | (0.66, 3.51) | 0.32 | 1.81 | (0.83, 3.97) | 0.14 |

Note: Model 1 adjusted for sex, age at serology, and age at Aβ-PET measurement. Model 2 adjusts for model 1 covariates plus APOE ε4 carriage. Model 3 adjusts for model 1 covariates plus education. The first tertiles of seroreactivity values are the reference categories

Abbreviations: HSV1 = Herpes simplex virus-1; HSV2 = Herpes simplex virus-2; VZV = Varicella zoster virus; EBV = Epstein-Barr virus; CMV = Cytomegalovirus; HHV6A = Human herpesvirus-6a; HHV6B = Human herpesvirus-6b; HHV7 = Human herpesvirus-7; BK = BK virus; JC = JC virus; MCV = Merkel cell polyomavirus.

**eFigure 1. Associations of pathogen antigen seroreactivities with plasma p-tau217 concentrations (at the median of the p-tau distribution).**

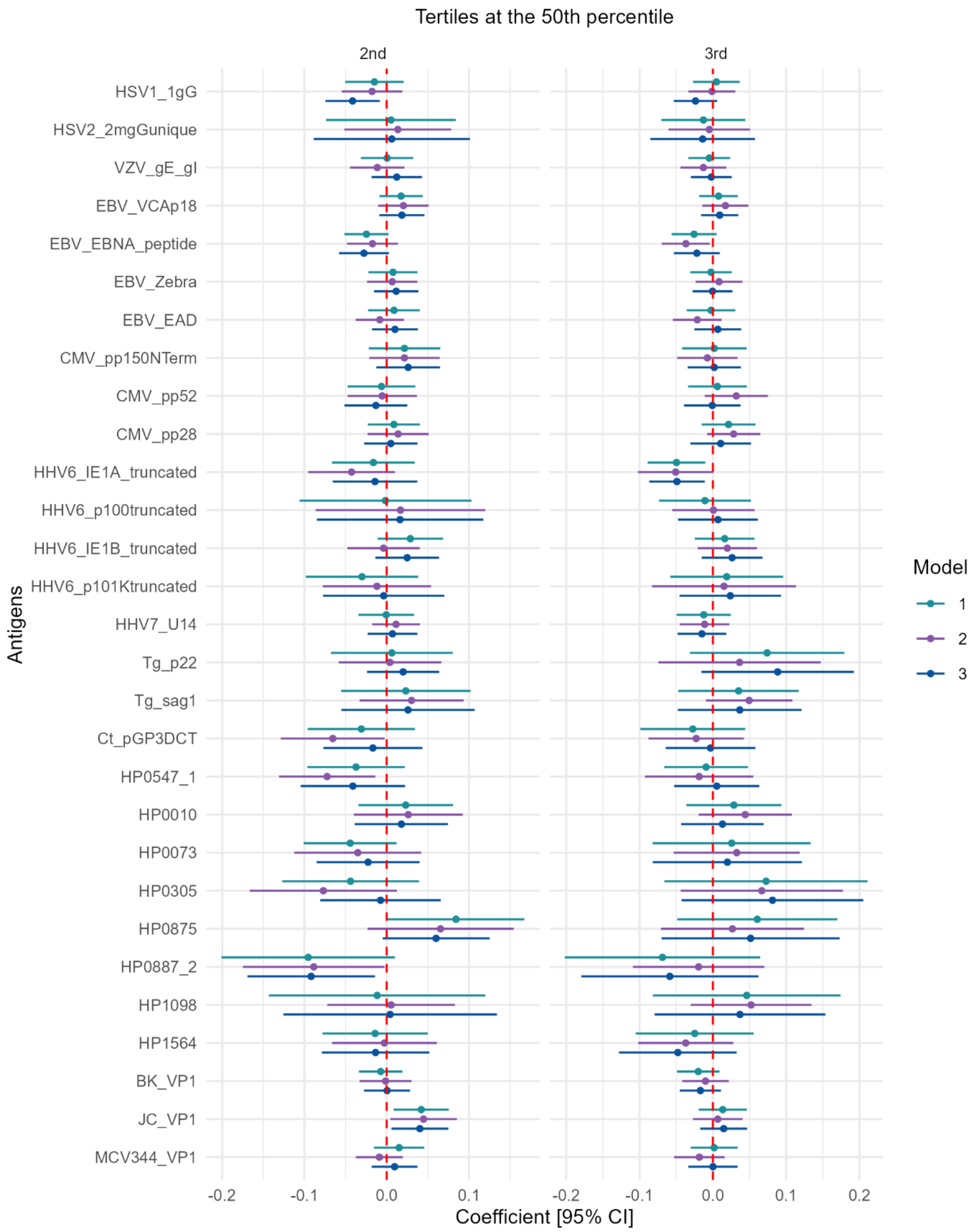

Forest plots show weighted quantile regression results depicting differences in p-tau217 concentrations at the 50$^{th}$ quantile (i.e. median) of the p-tau distribution in tertiles 2 (left panel) and 3 (right panel) of seroreactivity distributions, relative to individuals in tertile 1. Model 1 adjusted for sex, age at serology, and age at p-tau 217 measurement. Model 2 adjusts for model 1 covariates plus *APOE* ε4 carriage. Model 3 adjusts for model 1 covariates plus education.

Abbreviations: HSV1 = Herpes simplex virus-1; HSV2 = Herpes simplex virus-2; VZV = Varicella zoster virus; EBV = Epstein-Barr virus; CMV = Cytomegalovirus; HHV6A = Human herpesvirus-6a; HHV6B = Human herpesvirus-6b; HHV7 = Human herpesvirus-7; BK = BK virus; JC = JC virus; MCV = Merkel cell polyomavirus; Tg = *Toxoplasma gondii*; Hp = *Helicobacter pylori*; Ct = *Chlamydia trachomatis.*

**eFigure 2. Associations of pathogen antigen seroreactivities with plasma p-tau 217 concentrations (at the 75th quantile of the p-tau distribution).**

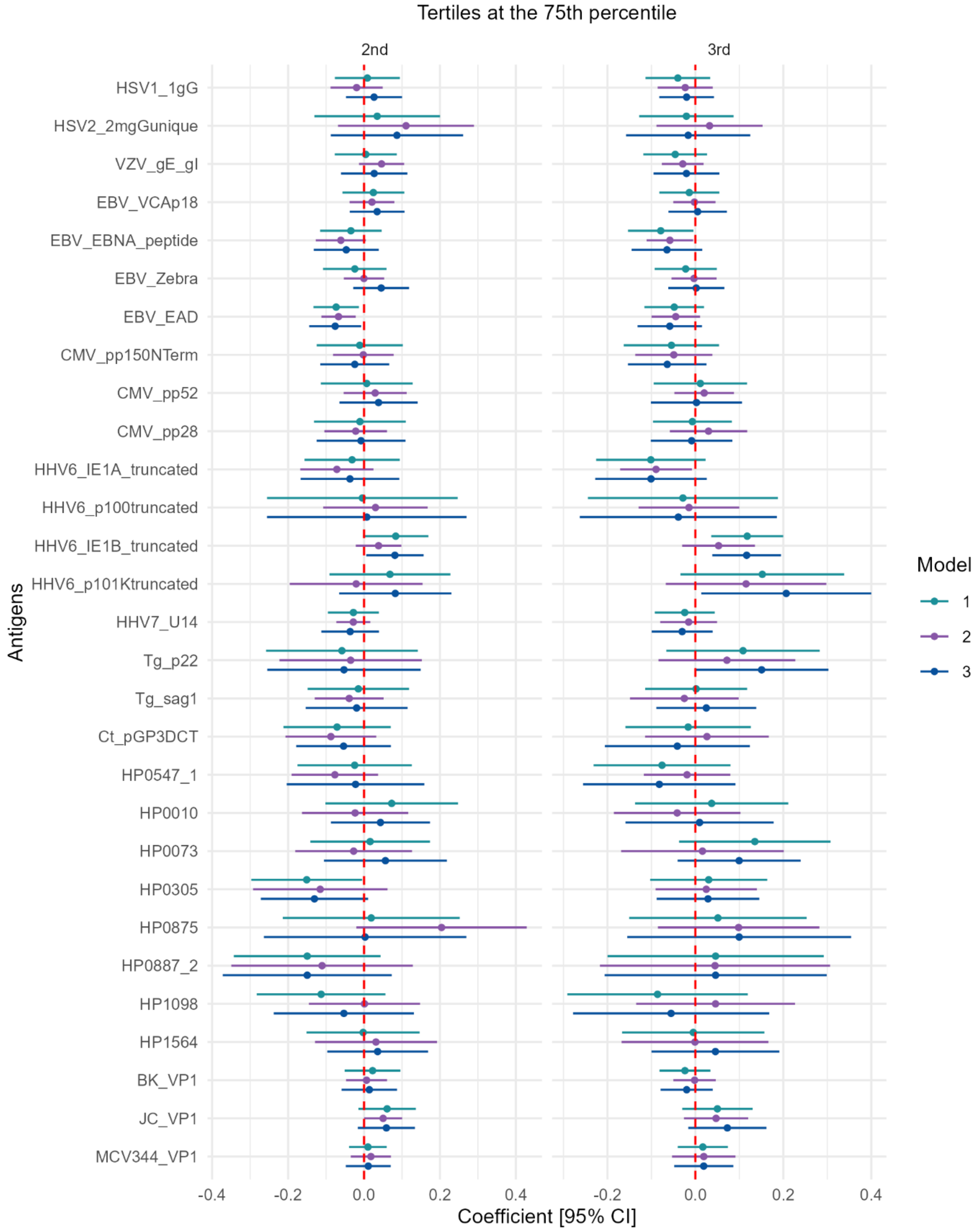

Forest plots show weighted quantile regression results depicting differences in p-tau 217 concentrations at the 75th quantile of the p-tau distribution in tertiles 2 (left panel) and 3 (right panel) of seroreactivity distributions, relative to individuals in tertile 1. Model 1 adjusted for sex, age at serology, and age at p-tau 217 measurement. Model 2 adjusts for model 1 covariates plus *APOE* ε4 carriage. Model 3 adjusts for model 1 covariates plus education.

Abbreviations: HSV1 = Herpes simplex virus-1; HSV2 = Herpes simplex virus-2; VZV = Varicella zoster virus; EBV = Epstein-Barr virus; CMV = Cytomegalovirus; HHV6A = Human herpesvirus-6a; HHV6B = Human herpesvirus-6b; HHV7 = Human herpesvirus-7; BK = BK virus; JC = JC virus; MCV = Merkel cell polyomavirus; Tg = *Toxoplasma gondii*; Hp = *Helicobacter pylori*; Ct = *Chlamydia trachomatis.*

**eFigure 3. Associations of pathogen serostatus and pathogen burden with odds of cerebral amyloidosis measured by Aβ-PET.**

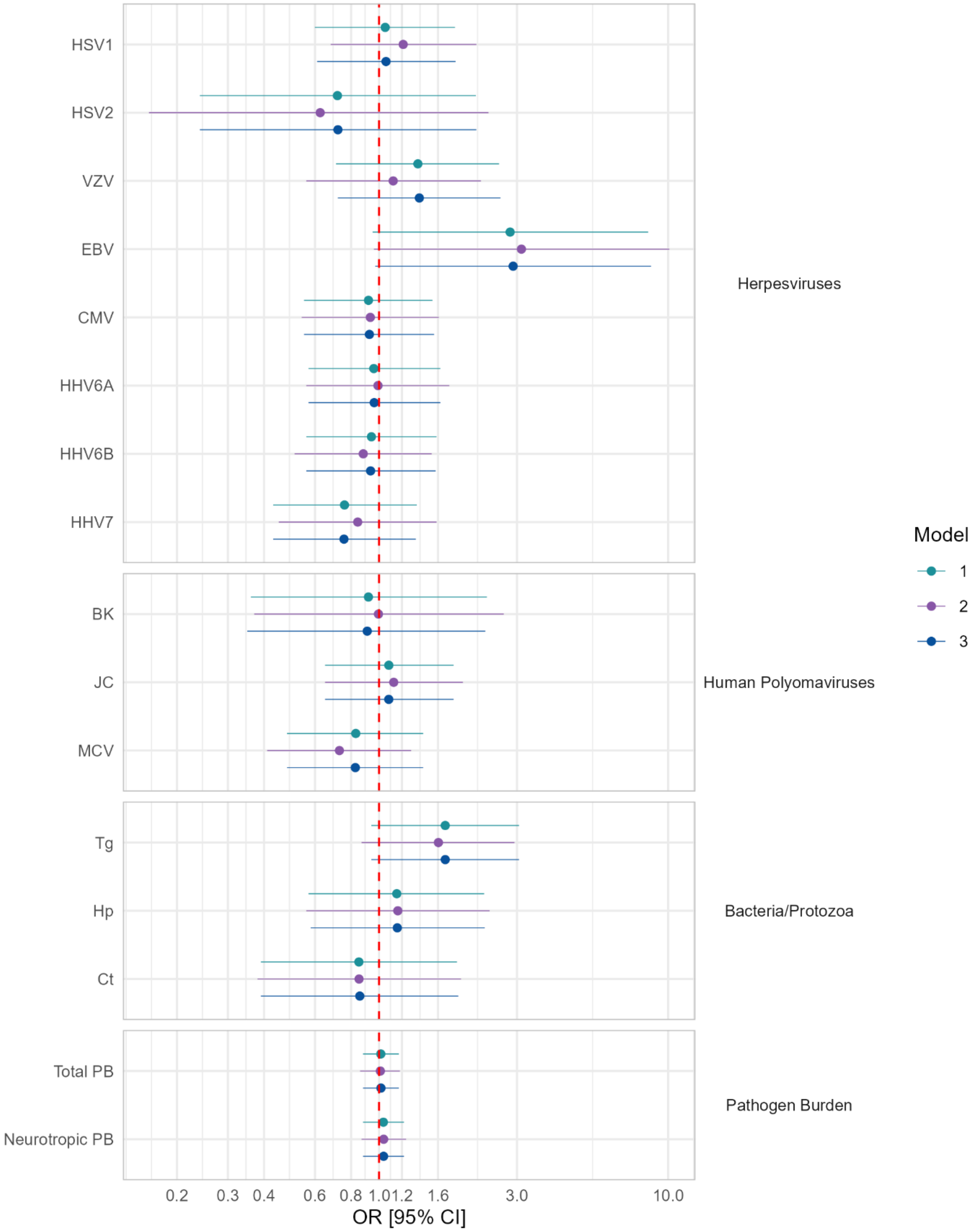

Forest plots show odds ratios (ORs) for a positive Aβ-PET scan per unit difference in the exposure – seropositive with reference to seronegative for individual pathogens, or per 1 additional pathogen in the pathogen burden indices. Model 1 adjusted for sex, age at serology, and age at Aβ-PET measurement. Model 2 adjusts for model 1 covariates plus *APOE* ε4 carriage. Model 3 adjusts for model 1 covariates plus education. A log in base 10 scale was applied to the x axis.

Abbreviations: HSV1 = Herpes simplex virus-1; HSV2 = Herpes simplex virus-2; VZV = Varicella zoster virus; EBV = Epstein-Barr virus; CMV = Cytomegalovirus; HHV6A = Human herpesvirus-6a; HHV6B = Human herpesvirus-6b; HHV7 = Human herpesvirus-7; BK = BK virus; JC = JC virus; MCV = Merkel cell polyomavirus; Tg = Toxoplasma gondii; Hp = Helicobacter pylori; Ct = Chlamydia trachomatis.

**eFigure 4. Associations of pathogen antigen seroreactivities with odds of cerebral amyloidosis measured by Aβ-PET.**

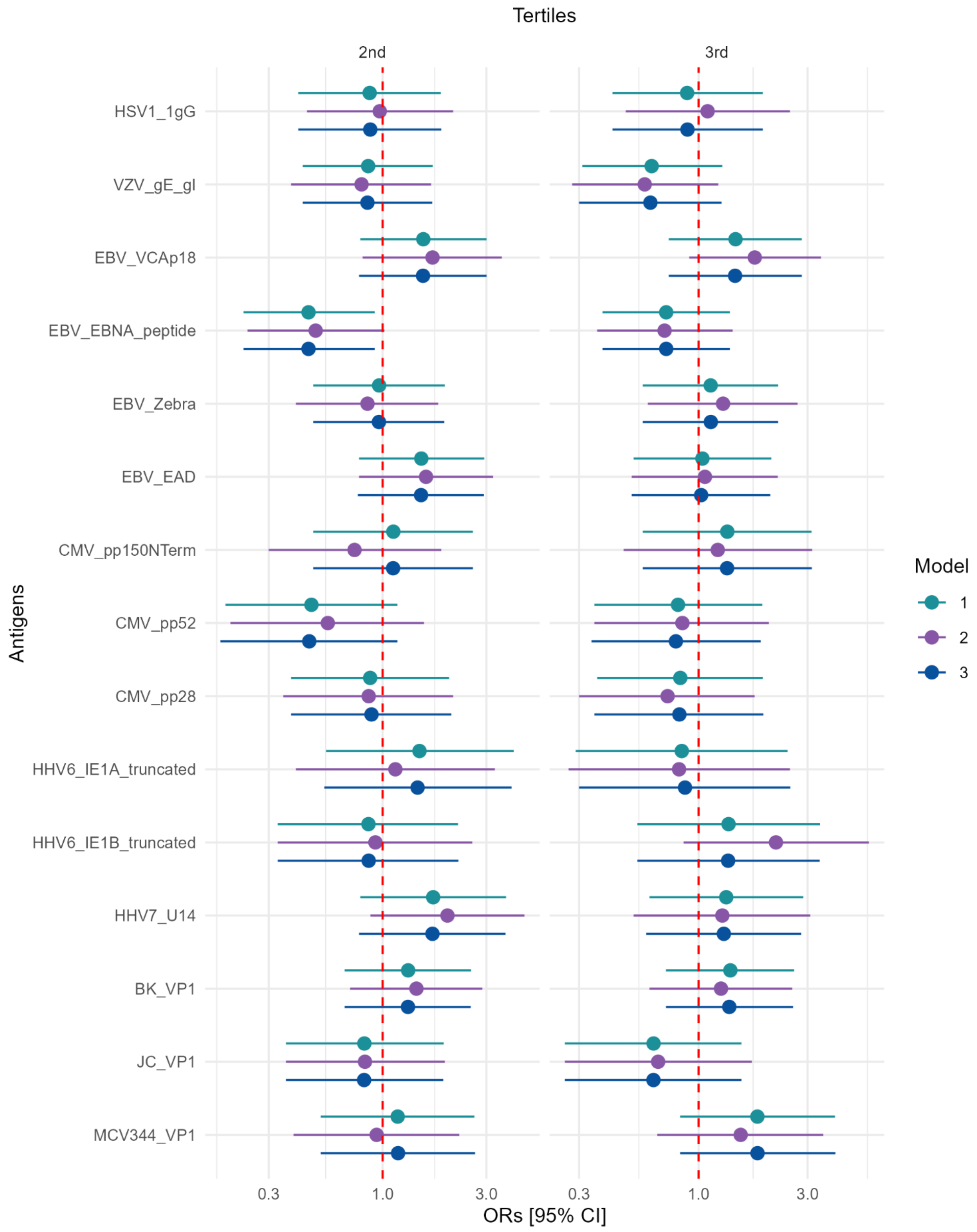

Forest plots show weighted odds ratios (ORs) for a positive Aβ-PET scan in tertiles 2 (left panel) and 3 (right panel) of seroreactivity distributions, relative to individuals in tertile 1. Model 1 adjusted for sex, age at serology, and age at Aβ-PET measurement. Model 2 adjusts for model 1 covariates plus *APOE* ε4 carriage. Model 3 adjusts for model 1 covariates plus education. A log in base 10 scale was applied to the x axis.

Abbreviations: HSV1 = Herpes simplex virus-1; HSV2 = Herpes simplex virus-2; VZV = Varicella zoster virus; EBV = Epstein-Barr virus; CMV = Cytomegalovirus; HHV6A = Human herpesvirus-6a; HHV6B = Human herpesvirus-6b; HHV7 = Human herpesvirus-7; BK = BK virus; JC = JC virus; MCV = Merkel cell polyomavirus.
